# Integrated Host/Microbe Metagenomics Enables Accurate Lower Respiratory Tract Infection Diagnosis in Critically Ill Children

**DOI:** 10.1101/2022.12.01.22282994

**Authors:** Eran Mick, Alexandra Tsitsiklis, Jack Kamm, Katrina L. Kalantar, Saharai Caldera, Amy Lyden, Michelle Tan, Angela M. Detweiler, Norma Neff, Christina M. Osborne, Kayla M. Williamson, Victoria Soesanto, Matthew Leroue, Aline B. Maddux, Eric A. F. Simões, Todd C. Carpenter, Brandie D. Wagner, Joseph L. DeRisi, Lilliam Ambroggio, Peter M. Mourani, Charles R. Langelier

## Abstract

**BACKGROUND:** Lower respiratory tract infection (LRTI) is a leading cause of death in children worldwide. LRTI diagnosis is challenging since non-infectious respiratory illnesses appear clinically similar and existing microbiologic tests are often falsely negative or detect incidentally-carried microbes, resulting in antimicrobial overuse and adverse outcomes. Lower airway metagenomics has the potential to detect host and microbial signatures of LRTI. Whether it can be applied at scale and in a pediatric population to enable improved diagnosis and treatment remains unclear.

**METHODS:** We used tracheal aspirate RNA-sequencing to profile host gene expression and respiratory microbiota in 261 children with acute respiratory failure. We developed a gene expression classifier for LRTI by training on patients with an established diagnosis of LRTI (n=117) or of non-infectious respiratory failure (n=50). We then developed a classifier that integrates the host LRTI probability, abundance of respiratory viruses, and dominance in the lung microbiome of bacteria/fungi considered pathogenic by a rules-based algorithm.

**RESULTS:** The host classifier achieved a median AUC of 0.967 by cross-validation, driven by activation markers of T cells, alveolar macrophages and the interferon response. The integrated classifier achieved a median AUC of 0.986 and increased the confidence of patient classifications. When applied to patients with an uncertain diagnosis (n=94), the integrated classifier indicated LRTI in 52% of cases and nominated likely causal pathogens in 98% of those

**CONCLUSIONS:** Lower airway metagenomics enables accurate LRTI diagnosis and pathogen identification in a heterogeneous cohort of critically ill children through integration of host, pathogen, and microbiome features.

## INTRODUCTION

Lower respiratory tract infection (LRTI) causes more deaths each year than any other type of infection and disproportionately impacts children(1–4). The ability to accurately determine whether LRTI underlies or contributes to respiratory failure in the intensive care unit and to identify the etiologic pathogens is critical for effective and targeted treatments. However, LRTI diagnosis is challenging since non-infectious respiratory conditions can appear clinically similar. Moreover, no microbiologic diagnosis is obtained in many cases of suspected LRTI since standard tests (such as bacterial culture) suffer from a narrow spectrum of targets and limited sensitivity (3, 5–8). At the same time, children are especially susceptible to false positive diagnoses due to frequent incidental carriage of potentially pathogenic microbes(3, 5, 9–12). As such, LRTI treatment is often empirical, leading to antimicrobial overuse, selection for resistant pathogens, and adverse outcomes(13–15).

Profiling host gene expression in the blood has shown promise as an innovative modality for diagnosing respiratory infection in hospitalized patients(16, 17). However, this approach has not been well studied in the diagnostically challenging critically ill pediatric population. Moreover, while blood gene expression can in some cases distinguish between the response to viral and bacterial infection(16–21), it cannot pinpoint the specific pathogens active in the respiratory tract, which is critical for optimal antimicrobial therapy.

Metagenomic next generation sequencing (mNGS) of lower airway samples (e.g., tracheal aspirate) has the potential to detect pathogens and host gene expression signatures of LRTI(22). Whether such an approach can be successfully applied at scale for the purpose of clinical diagnosis remains unclear. Its applicability in a pediatric population has also never been examined despite well-established age-related differences in LRTI epidemiology(3, 9), rates of incidental pathogen carriage(3, 5, 9), and the immune response to infection(23, 24). Furthermore, to our knowledge, no metagenomic approach for LRTI diagnosis thus far integrates host and microbial features into a single diagnostic output, a crucial step toward streamlined clinical application.

Here, we perform metagenomic RNA sequencing of tracheal aspirate in a prospective cohort of 261 children with acute respiratory failure requiring mechanical ventilation. We develop a host gene expression classifier for LRTI by training on patients with an LRTI diagnosis supported by clinical microbiologic testing and patients with respiratory failure due to non-infectious causes. We then develop a classifier that integrates host, pathogen, and microbiome features to accurately diagnose LRTI and identify the likely causal pathogens, including in cases with negative clinical microbiologic testing. Our results demonstrate the feasibility of lower airway metagenomics for improved LRTI diagnosis in a large and heterogeneous cohort and reveal the importance of profiling both the pulmonary immune response and microbiome in a pediatric population.

## RESULTS

### Patient cohort and LRTI adjudication

We enrolled children with acute respiratory failure requiring mechanical ventilation at eight hospitals in the United States between February 2015 and December 2017, as previously described(9, 25). Tracheal aspirate (TA) was collected within 24 hours of intubation and underwent metagenomic next generation sequencing (mNGS) of RNA to assay host gene expression and detect respiratory microbiota (**Figure 1**). High-quality host gene expression and microbial data was obtained for 261 patients.

**Figure 1:**
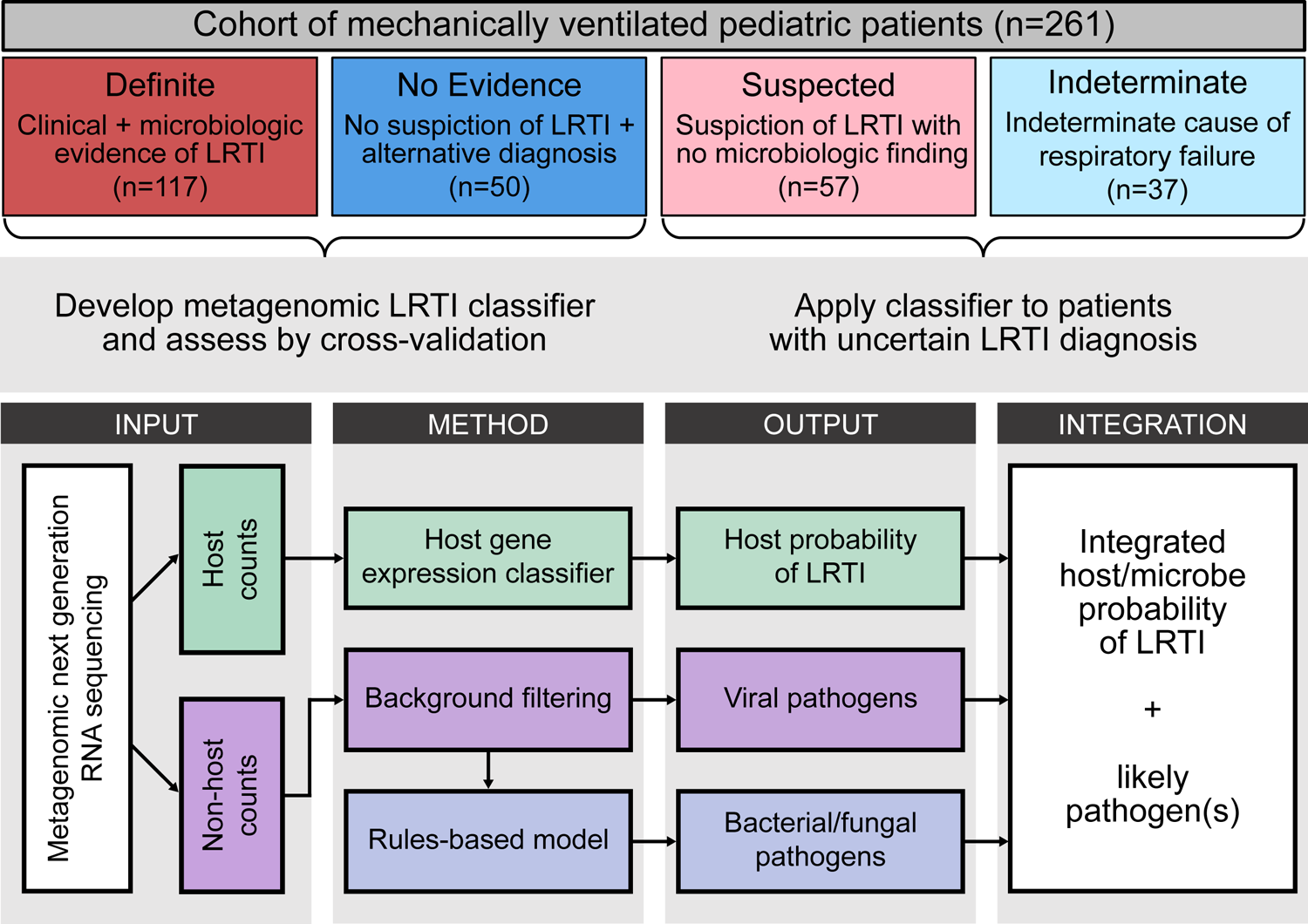
Study overview. Pediatric patients with acute respiratory failure requiring mechanical ventilation were clinically adjudicated into four LRTI status groups. The Definite and No Evidence groups, whose LRTI status was presumed to be known, were used to develop an integrated host/microbe mNGS classifier for LRTI and to evaluate its performance by cross-validation. The classifier was then applied to the Suspected and Indeterminate groups, whose LRTI status was considered uncertain. The integrated mNGS classifier combines a host probability of LRTI derived from the host gene counts, and features of any viral or bacterial/fungal pathogens derived from the non-host (microbial) taxon counts.

Adjudication of LRTI status was blinded to mNGS results and depended on the combination of two elements: i) a retrospective clinical diagnosis made by study-site clinicians, who reviewed all clinical, laboratory and imaging data available at the end of the admission, and ii) any standard-of-care respiratory microbiologic diagnostics performed during the admission (nasopharyngeal swab viral PCR and/or TA culture). Patients were assigned to one of four LRTI status groups, as follows: i) Definite, if clinicians made a diagnosis of LRTI and the patient had clinical microbiologic findings (n=117); ii) Suspected, if clinicians made a diagnosis of LRTI but there were no microbiologic findings (n=57); iii) Indeterminate, if no diagnosis of LRTI was made despite some microbiologic findings (n=37); and iv) No Evidence, if clinicians identified a clear non-infectious cause of acute respiratory failure and no clinical or microbiologic suspicion of LRTI arose (n=50) (**Figure 1**). We note that comprehensive microbiologic testing was not always performed in the No Evidence group in the absence of clinical suspicion.

The Definite and No Evidence groups were used to develop the metagenomic classifiers and to evaluate their performance by cross-validation due to the high degree of confidence in their clinical diagnoses (**Figure 1**). The patients in the Definite group were 39% female with a median age of 0.5 years (IQR 0.2-1.8) while the patients in the No Evidence group were 50% female with a median age of 6.5 years (IQR 1.5-12.9) (**Table 1**; **Supplemental Figure 1**). The difference in the age distribution of the two groups (p < 0.001, Mann-Whitney test) reflected recognized epidemiological distinctions in the conditions typically leading to respiratory failure in very young versus older children(3, 5).

**Table 1:** Demographic and clinical cohort characteristics.

|  | **Definite**<br>**(n=117)** | **No Evidence**<br>**(n=50)** | **P-value\*** | **Suspected**<br>**(n=57)** | **Indeterminate**<br>**(n=37)** |
| --- | --- | --- | --- | --- | --- |
| **Female, n (%)** | 45 (38.5%) | 25 (50.0%) | 0.18 | 26 (45.6%) | 16 (43.2%) |
| **Age, median [IQR]** | 0.5 [0.2, 1.8] | 6.5 [1.5, 12.9] | <0.001 | 1.7 [0.5, 6.0] | 1.5 [0.6, 10.8] |
| **Race, n (%)** |  |  |  |  |  |
| White | 69 (59.0%) | 30 (60.0%) | 0.99 | 33 (57.9%) | 20 (54.1%) |
| Black/African American | 26 (22.2%) | 7 (14.0%) | 0.29 | 11 (19.3%) | 10 (27.0%) |
| Asian | 5 (4.3%) | 6 (12.0%) | 0.088 | 2 (3.5%) | 2 (5.4%) |
| American Indian or Alaskan Native | 1 (0.9%) | 1 (2.0%) | 0.99 | 1 (1.8%) | 0 (0.0%) |
| Native Hawaiian/Other Pacific Islander | 1 (0.9%) | 0 (0.0%) | 0.99 | 0 (0.0%) | 0 (0.0%) |
| More than one race | 3 (2.6%) | 1 (2.0%) | 0.99 | 1 (1.8%) | 1 (0.03%) |
| Unknown | 12 (10.3%) | 5 (10.0%) | 0.99 | 9 (15.8%) | 4 (10.8%) |
| **Hispanic or Latino, n (%)** | 17 (14.5%) | 6 (12.0%) | 0.81 | 14 (24.6%) | 7 (18.9%) |
| **Comorbidities (CCC)†, n (%)** | 38 (32.5%) | 26 (52.0%) | 0.024 | 34 (59.7%) | 14 (37.8%) |
| **Immunosuppressed, n (%)** | 3 (2.6%) | 7 (14.0%) | 0.0085 | 5 (8.8%) | 6 (16.2%) |
| **Admission category, n (%)** |  |  |  |  |  |
| Medical | 117 (100.0%) | 28 (56.0%) | p<0.001 | 57 (100.0%) | 29 (78.4%) |
| Surgical | 0 (0.0%) | 15 (30.0%) | p<0.001 | 0 (0.0%) | 3 (8.1%) |
| Trauma | 0 (0.0%) | 7 (14.0%) | p<0.001 | 0 (0.0%) | 4 (10.8%) |
| **Time from hospital admission to intubation (hours), median [IQR]** | 4.8 [0.0, 23.6] | 3.5 [0.0, 20.9] | 0.60 | 2.6 [0.0, 15.9] | 1.0 [0.0, 26.5] |
| **Abx on or before sample date‡, n(%)** | 112 (95.7%) | 42 (84.0%) | 0.022 | 51 (89.5%) | 30 (69.8%) |

\*Nominal p-values comparing Definite and No Evidence patients. Mann-Whitney test used for all continuous variables. Fisher's exact test used for all categorical variables. †Complex Chronic Conditions(62). ‡Antibiotic treatment started on or prior to the date of sample collection.

Within the Definite group, 95% of patients were intubated by two days from hospital admission, indicative of community-acquired infection (**Table 1**). Clinical microbiologic testing identified viral infection alone in 46% of patients, bacterial infection alone in 14% of patients, and viral/bacterial co-infection in 40% of patients. The most common pathogens were respiratory syncytial virus (RSV) and *Haemophilus influenzae*, which frequently co-occurred(9). Diagnoses in the No Evidence group included trauma, neurological conditions, cardiovascular disease, airway abnormalities, ingestion of drugs/toxins, and sepsis that was clearly unconnected to LRTI. Nevertheless, most patients received antibiotic treatment by the time of TA sample collection in both the Definite (96%) and No Evidence (84%) groups (**Table 1**).

### Classification of LRTI status based on TA host gene expression features

We first compared TA host gene expression between the Definite and No Evidence groups to determine whether it could distinguish patients based on LRTI status, regardless of the underlying cause of infection. We identified 4,718 differentially expressed genes at a Benjamini-Hochberg adjusted p < 0.05 (**Supplemental Figure 2A**; **Supplemental Data 2**). As expected, gene set enrichment analysis identified elevated expression of pathways involved in the immune response to infection in the Definite group (**Supplemental Figure 2B**; **Supplemental Data 3**). Pathways related to the interferon response, a hallmark of anti-viral innate immunity, were most strongly upregulated, consistent with the high prevalence of viral infections in the Definite group. Additional immune pathways upregulated in this group included toll-like receptor signaling, cytokine signaling, inflammasome activation, neutrophil degranulation, antigen processing, and B cell and T cell receptor signaling. Conversely, pathways with reduced expression in the Definite group included translation, cilium assembly and lipid metabolism (**Supplemental Figure 2B**; **Supplemental Data 3**).

Since we observed a clear host signature of infection, we developed a classification approach to distinguish the Definite and No Evidence patients based on gene expression and evaluated its performance by 5-fold cross-validation. For each train/test split, we: i) used lasso logistic regression on the samples in the training folds to select a parsimonious set of informative genes, ii) trained a random forest classifier using the selected genes, and iii) applied it to the samples in the test fold to obtain a host probability of LRTI.

Our approach yielded a median area under the receiver operating characteristic curve (AUC) of 0.967 (range: 0.953-0.996), with the number of genes selected for use in the classifier ranging from 11 to 25 across the five train/test splits (**Figure 2A**; **Supplemental Table 1**). Using a 50% out-of-fold probability threshold to classify a patient as suffering from LRTI (LRTI+), the classifier assigned 92% of Definite patients and 80% of No Evidence patients according to their clinical LRTI adjudication (**Figure 2B**).

**Figure 2:**
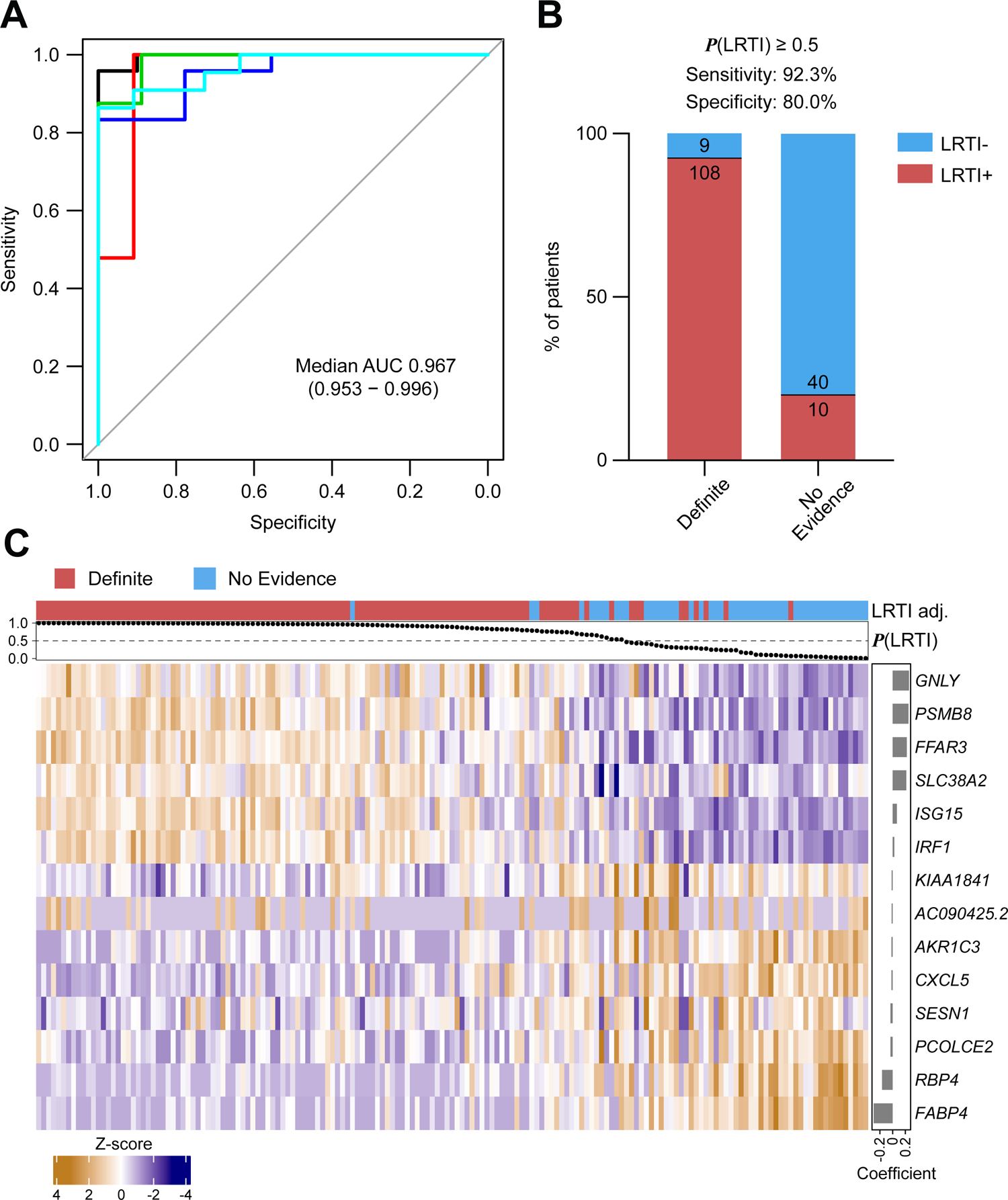
Host gene expression classifier for LRTI diagnosis. **A)** Receiver operating characteristic (ROC) curve of the host gene expression classifier in each of the test folds. The median and range of the area under the curve (AUC) are indicated. **B)**Bar plot showing the number and percentage of Definite and No Evidence patients that were classified according to their clinical adjudication using a 50% out-of-fold probability threshold. **C)** Heatmap showing standardized variance-stabilized expression values across all patients (columns) for the 14 final classifier genes (rows) selected from the full Definite and No Evidence dataset. Shown are the LRTI adjudication (top colored horizontal bar) and out-of-fold LRTI probability (top dot plot) of each patient, and the regression coefficient of each selected gene (side bar plot).

Having validated the performance of our approach by cross-validation, we then applied lasso logistic regression to all the Definite and No Evidence patients to select a final set of genes (n=14) for later classification of patients with Suspected or Indeterminate LRTI status (**Figure 2C**; **Supplemental Table 2**). As expected, the genes in the final classifier set that were assigned high absolute regression coefficients were also repeatedly selected in the cross-validation procedure (**Supplemental Table 2**).

The selected genes with the most positive regression coefficients, corresponding to higher expression in the Definite group, were: *GNLY*, encoding an anti-bacterial peptide present in cytolytic granules of cytotoxic T cells and natural killer cells(26); *SLC38A2*, encoding a glutamine transporter upregulated in CD28-stimulated T cells(27, 28); *FFAR3*, encoding a G protein-coupled receptor activated by short-chain fatty acids that is induced by alveolar macrophages upon infection(29); and the interferon-stimulated genes *PSMB8*, *ISG15* and *IRF1* (**Figure 2C**; **Supplemental Table 2**).

The selected genes with the most negative regression coefficients, corresponding to lower expression in the Definite group, were: *FABP4*, encoding a fatty acid-binding protein considered a marker of alveolar macrophages, whose expression in the lung decreases in patients with LRTI, including COVID-19 (30–32); and *RBP4*, encoding a retinol-binding protein, whose expression in the lung has also been shown to sharply decrease following onset of LRTI(30) and whose expression by macrophages in vitro is depressed by inflammatory stimuli(33) (**Figure 2C**; **Supplemental Table 2**).

We examined the expression of the final classifier genes as a function of patient age to confirm that their selection was not influenced by the different age distributions of the Definite and No Evidence groups (**Supplemental Figure 3**). Reassuringly, we found no significant difference in the expression of the 14 genes when comparing No Evidence patients under the age of four (n=23; median age 1.3 years) and over the age of four (n=27; median age 12.5) (**Supplemental Table 3A**). Further, we found that expression of 12 of the genes remained significantly different when comparing only children under the age of four in the Definite (n=100; median age 0.4) and No Evidence (n=23; median age 1.3) groups (**Supplemental Table 3B**).

### Detection of pathogens by mNGS and definition of microbial classification features

We proceeded to analyze the microbial mNGS data to nominate likely pathogens whose features could be integrated into the LRTI classifier to increase confidence in the results and whose identity could be used to guide treatment. We processed the TA samples alongside water controls through the CZ-ID metagenomic analysis pipeline to obtain a count matrix of microbial taxa. The water controls allowed us to generate a background count distribution for each taxon, which modeled the contribution of contamination by microbes present in the laboratory environment or reagents.

Viruses with known ability to cause LRTI that were present at an abundance statistically exceeding their background distribution were considered probable pathogens. By this criterion, we detected viruses in the lungs of 107/117 (91%) Definite patients, with RSV the most prevalent (**Figure 3A**). Among No Evidence patients, 8/50 (16%) also had viruses detected by mNGS, which were probably missed clinically in the absence of characteristic symptoms. We defined the summed abundance of all pathogenic viruses detected in a patient, measured in reads-per-million (rpM), as the patient’s ‘viral score’ for later use in an integrated host/microbe classifier (**Figure 3B**).

**Figure 3:**
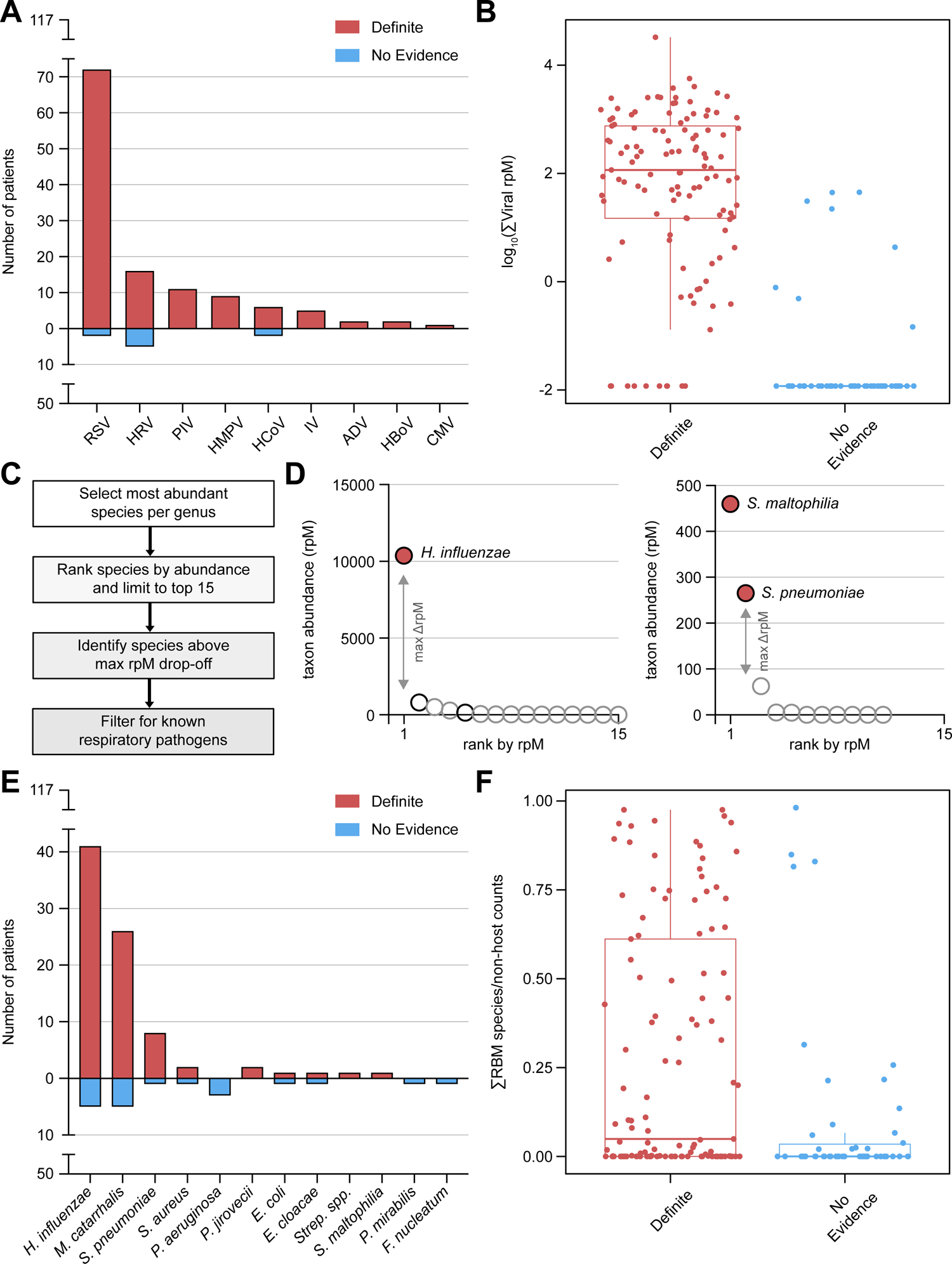
Metagenomic identification of respiratory pathogens. **A)** Bar plot showing the distribution of viruses detected by mNGS after background filtering in the Definite and No Evidence patients. RSV, respiratory syncytial virus; HRV, human rhinovirus; PIV, parainfluenza virus; HMPV, human metapneumovirus; HCoV, human coronavirus; IV, influenza virus; ADV, adenovirus; HBoV, human bocavirus; CMV, cytomegalovirus. **B)** Boxplot showing the log_10_-transformed summed abundance, measured in reads-per-million (rpM), of all pathogenic viruses detected in each patient, separated by group. Prior to log_10_-transformation, the minimum non-zero rpM value in the dataset was divided by 10 and added to all the samples. Horizontal lines denote the median, box hinges represent the interquartile range (IQR), and whiskers extend to the most extreme value no greater than 1.5*IQR from the hinges. **C)** Analysis steps applied as part of the rules-based model (RBM), a heuristic approach designed to identify potential bacterial/fungal pathogens in the context of LRTI. **D)** Graphical illustration of the RBM results in two representative Definite patients. Each dot is a bacterial/fungal species most abundant in its respective genus. A species above the maximum drop-off in rpM has a red fill, otherwise the fill is white. A species on the list of known respiratory pathogens has a black outline, otherwise the outline is gray. **E)** Bar plot showing the distribution of bacteria/fungi called as potential pathogens by the RBM in the Definite and No Evidence patients. *Strep. spp.*, *Streptococcus* species other than *S. pneumoniae*. **F)** Boxplot showing the proportion of the RBM-identified pathogen(s) out of all non-host counts in each patient, separated by group. Horizontal lines denote the median, box hinges represent the interquartile range (IQR), and whiskers extend to the most extreme value no greater than 1.5*IQR from the hinges.

Because most Definite patients had a positive nasopharyngeal (NP) swab viral PCR test, we could compare the viruses detected by PCR and mNGS (**Supplemental Data 4**). The comparison was complicated, however, by the fact that PCR was performed on upper airway samples, so a virus detected by PCR was not necessarily present in the lower airway. Bearing this in mind, we found that 99/101 (98%) Definite patients with a viral PCR hit also had a virus detected by mNGS, and both approaches detected at least one virus in common in 91 (92%) of those patients (**Supplemental Figure 4A**). Most cases where NP swab PCR detected a virus, but mNGS did not, involved adenovirus (**Supplemental Figure 4B**). mNGS alone detected viruses in 8/16 (50%) Definite patients lacking a viral PCR hit (**Supplemental Figure 4A**). In a subset of Definite patients where we performed viral PCR on the same TA samples subjected to mNGS (n=21), 96% of PCR hits were detected by mNGS (**Supplemental Table 4**).

Bacterial and fungal taxa in the mNGS data also underwent background filtering to retain only those present at an abundance statistically exceeding their background distribution based on water controls. Because incidental carriage of potentially pathogenic bacteria is common in children, we additionally applied a previously published algorithm to distinguish possible pathogens from commensals, called the rules-based model (RBM)(9, 22). The RBM identifies bacteria and fungi with known pathogenic potential that are relatively dominant in a sample (**Figure 3, C and D**), based on the principle that uncontrolled growth of a pathogen leads to reduced lung microbiome alpha diversity in the context of LRTI(22, 34–36) (**Supplemental Figure 4, C and D**).

The RBM identified possible bacterial/fungal pathogens in 78/117 (66%) Definite patients, with the most common being *H. influenzae*, *Moraxella catarrhalis* and *Streptococcus pneumoniae* (**Figure 3E**). The RBM also identified potential bacterial/fungal pathogens in 17/50 (34%) No Evidence patients. Patients in the Definite group with an RBM-identified pathogen exhibited markedly lower bacterial alpha diversity compared to Definite patients without an RBM-identified pathogen and compared to No Evidence patients (**Supplemental Figure 4D**). In contrast, No Evidence patients with an RBM-identified pathogen did not typically exhibit a loss of bacterial alpha diversity (**Supplemental Figure 4D**), and in such cases the RBM-identified species was far less dominant (**Figure 3F**). We therefore defined the patient’s ‘bacterial score’ for use in an integrated host/microbe classifier as the proportion of the RBM-identified pathogens out of all non-host counts, a measure of relative dominance (**Figure 3F**).

We next sought to compare the bacterial and fungal pathogens identified by mNGS with those found by culture of TA samples in the Definite patients (**Supplemental Data 4**). Importantly, mNGS can detect organisms that are challenging to grow in culture or are inhibited by previous antibiotic treatment, and the RBM selects the likeliest pathogen based on a global view of the microbiome. Despite these inherent differences between culture and the RBM, we found that in 44/63 (70%) Definite patients who had a positive culture, at least one pathogen identified by the RBM was also found by culture (**Supplemental Figure 4E**). In the remaining 19 patients, the RBM identified a different species than culture (n=7), or no pathogen at all (n=12). Even in these cases, the species grown in culture was usually present in the mNGS data, but other species were more dominant (**Supplemental Figure 4E**). The RBM also identified a potential pathogen in 27/54 (50%) Definite patients lacking a positive culture (**Supplemental Figure 4E**). Most cases where the species grown in culture was absent from the mNGS data after background filtering involved *Staphylococcus aureus*, *Streptococcus* species other than *S. pneumoniae*, and *Escherichia coli* (**Supplemental Figure 4F**).

### Host gene expression differences between viral and bacterial LRTI

Overall, mNGS identified viral and/or bacterial pathogens in 114/117 (97%) Definite patients. Having established by mNGS which Definite patients had an exclusively bacterial infection (n=7), an exclusively viral infection (n=36), or a viral/bacterial co-infection (n=71), we went back and examined how effectively the top host classifier genes captured these different scenarios (**Supplemental Figure 5A**). As expected, some of the interferon-stimulated genes (e.g., *ISG15*) provided much more discriminating power for Definite patients with a viral infection as compared to those with a purely bacterial infection. Reassuringly, however, several other classifier genes behaved similarly regardless of the underlying infection type.

We then asked more broadly whether host gene expression differed between patients with any bacterial LRTI (including viral co-infection) and patients with purely viral LRTI. We identified 108 differentially expressed genes at a Benjamini-Hochberg adjusted p < 0.05 (**Supplemental Figure 5B**; **Supplemental Data 2**), and found that genes related to neutrophil degranulation and cytokine signaling were enriched in patients with any bacterial LRTI (**Supplemental Figure 5C**; **Supplemental Data 3**). These results suggest the potential for developing in future work a rule-out classifier for bacterial infection that could be used to limit unnecessary antibiotic usage.

### Classification of LRTI status based on integration of host and microbial features

Next, we asked whether integrating the host and microbial features could improve the performance of metagenomic LRTI classification. We fit a logistic regression model on the following three features: i) the LRTI probability output of the host classifier, ii) the summed abundance, measured in reads-per-million (rpM), of any pathogenic viruses present after background filtering (‘viral score’), and iii) the proportion of the potentially pathogenic bacteria/fungi identified by the RBM out of all non-host read counts (‘bacterial score’) (**Figure 4A**). As expected, the host and microbial features were correlated across most samples, but some notable exceptions were observed (**Supplemental Figure 6**).

**Figure 4:**
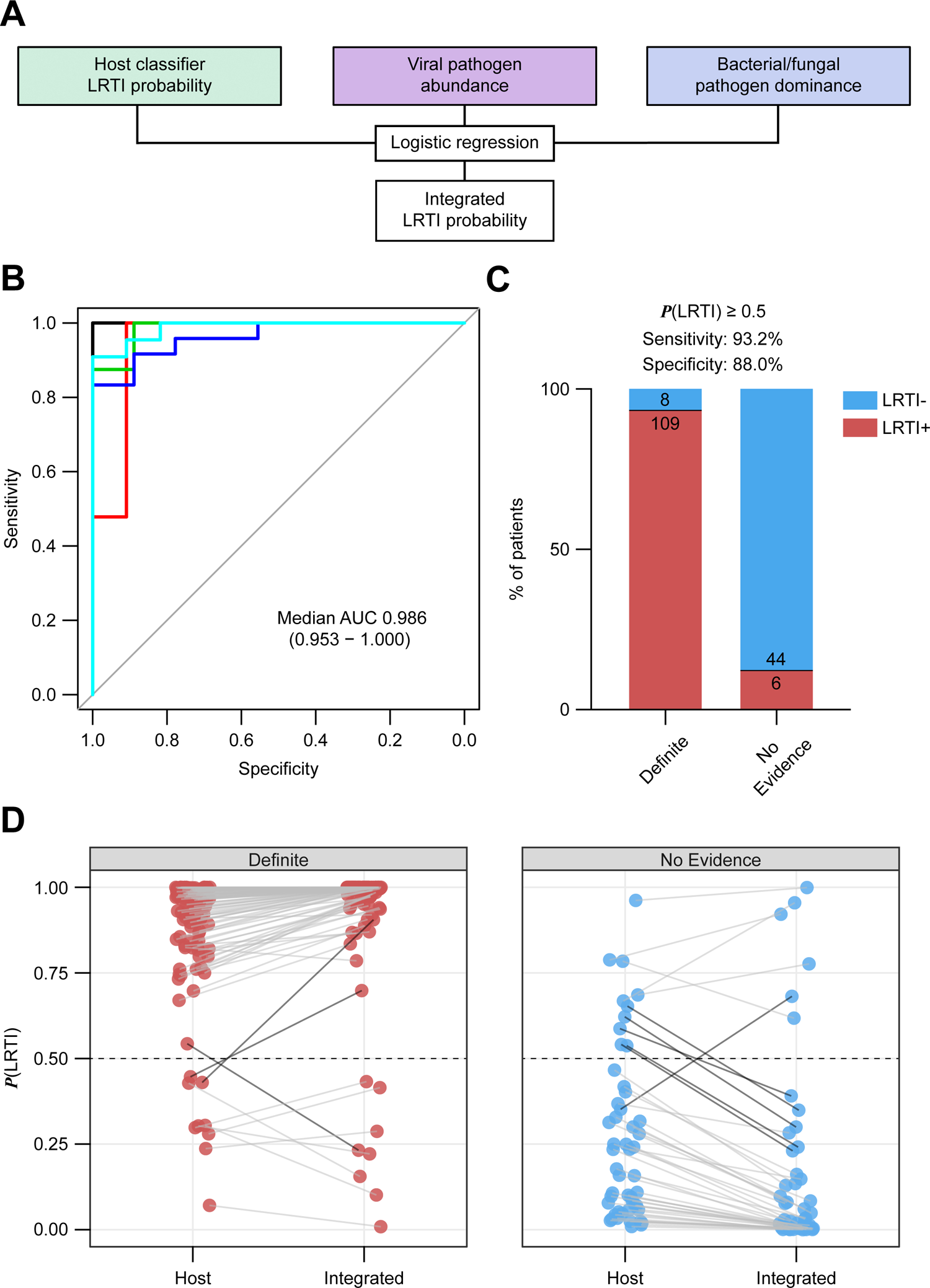
Integrated host/microbe classifier for LRTI diagnosis. **A)** Schematic of the integrated host/microbe classifier. **B)** Receiver operating characteristic (ROC) curve of the integrated classifier in each of the test folds. The median and range of the area under the curve (AUC) are indicated. **C)** Bar plot showing the number and percentage of Definite and No Evidence patients that were classified according to their clinical adjudication using a 50% out-of-fold probability threshold. **D)** The shift in out-of-fold LRTI probability from the host classifier to the integrated classifier for Definite (left panel) and No Evidence (right panel) patients. Dark connecting lines represent samples whose LRTI probability shifted across the 50% threshold.

The integrated classifier achieved an AUC of 0.986 (range: 0.953-1.000) when assessed by 5-fold cross-validation (**Figure 4B**; **Supplemental Table 5**), applying the same train/test splits used in the host-only cross-validation. Using an out-of-fold probability threshold of 50%, the integrated classifier assigned 109/117 (93%) Definite patients as LRTI+ and 44/50 (88%) No Evidence patients as LRTI-(**Figure 4C**; **Supplemental Table 6**). Compared to the host-only classifier, a net of five additional patients were now classified according to their clinical adjudication and the confidence of patient classifications increased, as reflected by more extreme output probabilities (**Figure 4D**). We note that at a much lower out-of-fold probability threshold of 15%, the integrated classifier’s sensitivity for LRTI in the Definite group rose to >98%, suggesting a use-case as a rule-out test for LRTI.

Finally, we trained the integrated host/microbe classifier on all the Definite and No Evidence patients and then applied it to the Suspected and Indeterminate patients, whose clinical diagnosis was less certain. The integrated classifier indicated 37/57 (65%) Suspected patients were LRTI+ compared with 12/37 (32%) Indeterminate patients (**Figure 5A**), consistent with the stronger clinical suspicion of LRTI in the former group. Across all 52 patients classified as LRTI+ in these two groups, likely pathogens (viral, bacterial, or fungal) were identified in 51 patients (98%). Pathogens detected included common (e.g., rhinovirus, *H. influenzae*), uncommon (e.g., bocavirus, parechovirus), and difficult to culture (e.g., *Mycoplasma pneumoniae*) microbes (**Figure 5B**). We also designed a visual summary incorporating all three inputs of the integrated classifier and its output LRTI probability (**Figure 5C**).

**Figure 5:**
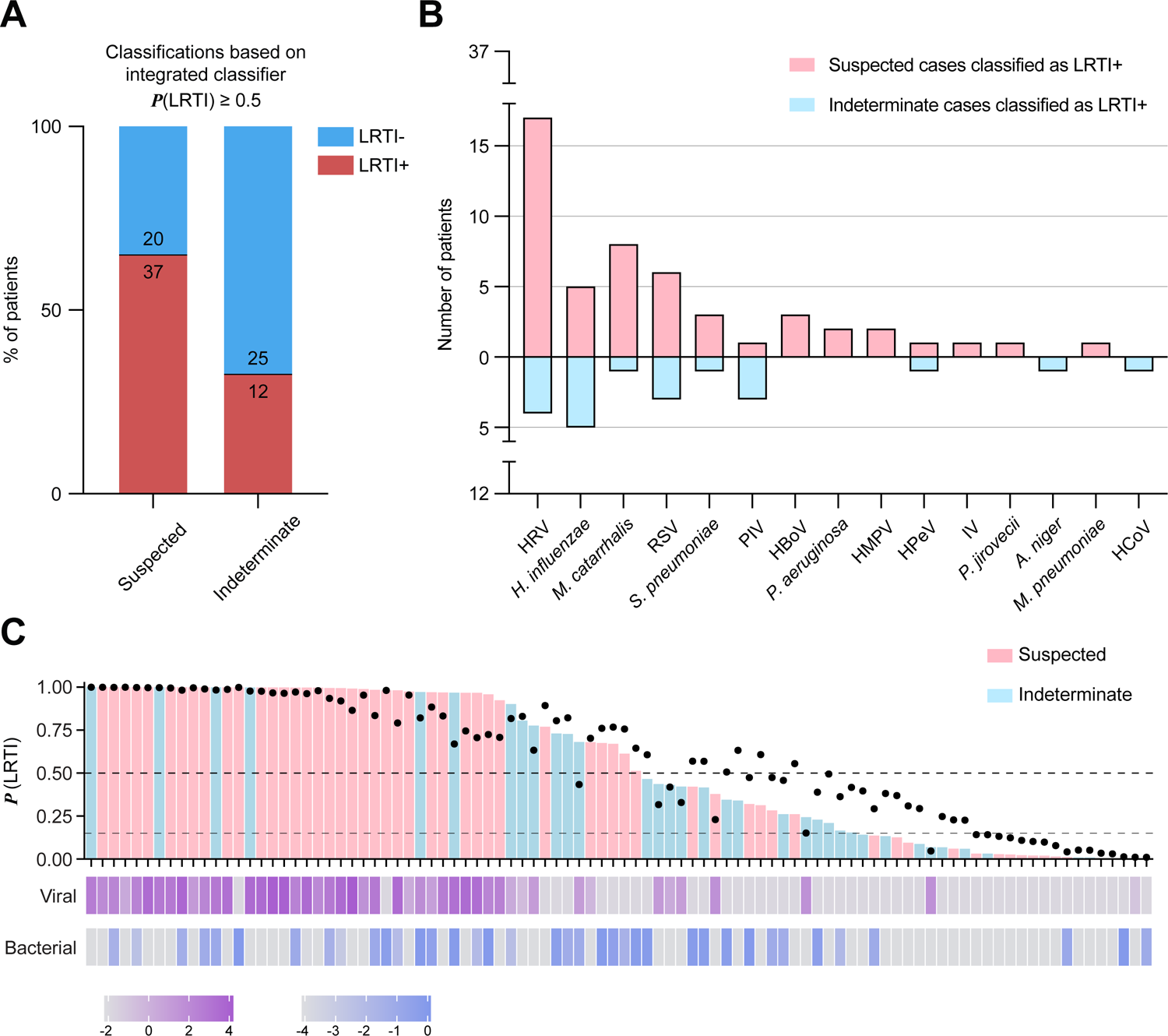
Application of the integrated classifier to Suspected and Indeterminate patients. **A)** Bar plot showing the number and percentage of Suspected and Indeterminate patients that were classified as LRTI+ by the integrated classifier using a 50% probability threshold. **B)** Viruses detected by mNGS and bacteria/fungi identified by the RBM across the patients classified as LRTI+ in the Suspected and Indeterminate groups. HRV, human rhinovirus; RSV, respiratory syncytial virus; PIV, parainfluenza virus; HBoV, human bocavirus; HMPV, human metapneumovirus; HPeV, human parechovirus; IV, influenza virus; HCoV, human coronavirus. **C)** Overview of inputs and output of the integrated classifier for all Suspected and Indeterminate patients. Top bars denote the integrated probability of LRTI and are colored by patient group; black dots represent the input host LRTI probability; bottom vertical bars show the input log_10_-transformed viral and bacterial scores. Dashed lines indicate the 50% LRTI probability threshold and the 15% rule-out threshold.

## DISCUSSION

Lower respiratory tract infection (LRTI) involves a dynamic relationship between pathogen, lung microbiome and host response that is not captured by existing clinical diagnostic tests. Here, we demonstrate that mNGS of lower respiratory samples enables accurate LRTI diagnosis based on features of each of these key elements in critically ill children, a demographic facing a high burden of LRTI. We build on proof-of-concept work in adults(22) to develop the first fully integrated host/microbe LRTI diagnostic classifier, and validate its performance in a large, multicenter prospective cohort.

Incidental carriage of pathogens in the respiratory tract is common in children(3, 5, 9–12). Consistent with this, detection of a pathogen by mNGS was in many cases insufficient for accurate LRTI diagnosis in our cohort. Among No Evidence patients, 40% had potentially pathogenic microbes detected by mNGS even after application of the RBM (for bacteria and fungi). This is notably different from adults, for whom both clinical and metagenomic studies have demonstrated much lower rates of incidental pathogen carriage(7, 22). Profiling the host response is thus particularly important for pediatric LRTI diagnosis, as it provides evidence of an immune response to infection.

Remarkably, an LRTI diagnostic classifier based on host gene expression performed very well on its own, with a median AUC of 0.967 by cross-validation. The host signature was driven by activation markers of T cells, alveolar macrophages, and the interferon response, and successfully captured cases of viral infection, bacterial infection, or co-infection. This performance suggests the gene signature could be incorporated into a clinical PCR assay as a standalone rapid diagnostic. It is likely that an even more parsimonious signature than the one used in the mNGS classifier would suffice, as six genes exhibited the most discriminating power.

The integrated host/microbe classifier achieved a median AUC of 0.986 by cross-validation. The incorporation of microbial features increased the confidence of LRTI classification, even though relatively few patients switched their assigned diagnosis. It is likely that the integrated classification approach will prove even more valuable in settings where the host signature may not perform as well on its own (e.g., immune-compromised patients), and will generalize better to future cohorts. Moreover, it provides clinicians with a unified framework both for LRTI diagnosis and etiologic pathogen identification.

Unlike for host gene expression, the microbial features in the integrated classifier were not automatically selected by training on identified taxa and their features in the Definite and No Evidence groups. Such an approach was not feasible given the sparse presence of individual respiratory pathogens across patients in the cohort, especially in the No Evidence group. As larger datasets are generated, it may be possible to use machine learning approaches to capture the ‘null distribution’ of incidentally carried pathogens in the lower respiratory tract and identify outlier cases that signal LRTI. Even then, designating a specific microbe as a ‘true’ causal pathogen for training purposes would be non-trivial, especially in cases of co-infection. Instead, we defined summary viral and bacterial scores motivated by accumulated clinical and microbiologic knowledge. For bacteria and fungi, we took advantage of the collapse of lung microbiome diversity in the setting of pathogen dominance, an established feature of LRTI(22, 34, 35).

Comparison of mNGS and clinical microbiologic testing was complicated by inherent differences in the anatomical site of testing (upper respiratory viral PCR vs. lower respiratory mNGS) or the question addressed (growth in culture vs. dominance by mNGS), as well as by heterogeneity in microbiologic practices among study sites. Nevertheless, when clinical testing identified a microbe, it was in most cases present in the mNGS data. A notable exception was adenovirus, which was consistently absent by mNGS when detected by NP swab PCR. This could reflect sensitivity limitations of RNA-sequencing for a DNA virus or true absence in the lower airway. Our secondary analysis revealing higher concordance of PCR and mNGS when performed on the same lower respiratory specimens, however, argues for the latter possibility. Future work could examine targeted enrichment strategies(37, 38) to improve detection of this or any other pathogen that proves challenging to capture by mNGS. Regardless, our findings highlight the value of concomitant assessment of the host response, which can accurately inform LRTI status even when pathogens are not detected.

A key advantage of mNGS is the capacity to provide a microbiologic diagnosis when traditional clinical testing returns negative, as in an estimated 20-60% of suspected community- or hospital-acquired pneumonia cases(3, 6–8). Indeed, the integrated mNGS classifier confirmed LRTI in 65% of children with suspected infection but negative clinician-ordered testing in our cohort, and in 32% of patients with respiratory failure of indeterminate etiology. It also provided a microbiologic diagnosis in all but one of these patients, highlighting the potential to inform pathogen-targeted versus empirical treatment.

Acute respiratory illnesses are a leading contributor to inappropriate antimicrobial use, a practice driven by challenges distinguishing LRTI from non-infectious causes of respiratory failure or distinguishing bacterial from viral LRTI. Reflecting this is the observation that 90% of children in our cohort received empiric antimicrobials by the time of sample collection, including 84% in the No Evidence group. Host/microbe mNGS offers an opportunity for improved antimicrobial stewardship, particularly in clinically uncertain cases, by providing a probability of infection and by nominating the likely pathogen. In fact, we found that the integrated classifier could be tuned to achieve >98% sensitivity for LRTI detection, highlighting its potential use as a rule-out test to help exclude the need for antimicrobials. Moreover, our host gene expression analysis revealed potential for development of a host classifier specifically for bacterial infection.

Our study has several limitations that should be kept in mind. In developing the mNGS classifiers, we relied on retrospective clinical adjudication for designating the ‘ground truth’ LRTI status of patients in the cohort. Retrospective adjudication, which considers the context of patient trajectory and clinical data not available at the time of initial admission, was the only practical approach. However, by nature, it is not infallible and was subject to variability in clinical and microbiologic practices across study sites and to the known limitations of standard microbiologic diagnostics. Moreover, comprehensive microbiologic testing was not always performed in the No Evidence group in the absence of clinical suspicion of LRTI, which likely allowed a few patients into this group who were suffering from unrecognized infection on top of their primary diagnosis. It is thus likely that some No Evidence patients deemed LRTI+ by the mNGS classifier were not truly misclassified, but rather incorrectly adjudicated. Study limitations also include the different age distributions of comparator groups and the relative paucity of purely bacterial infections.

mNGS provides a broad screen for bacteria, viruses, and other pathogens to overcome the limitations of traditional clinical microbiologic tests. Assays utilizing this technique are already in use in hospitals for microbe detection in typically sterile compartments, such as blood (sepsis) and cerebrospinal fluid (meningitis), with turnaround of ≤48 hours(39, 40). mNGS promises to improve the diagnosis and treatment of respiratory infections as well(9, 22, 41–45), but has not yet seen clinical translation in this area. Respiratory samples present a special challenge since they harbor microbial communities, including potential pathogens, even in states of health. Host gene expression can help distinguish bona fide infection, and several studies have demonstrated the utility of blood transcriptional profiling for this purpose(16, 17, 20). However, this approach precludes identification of the etiologic respiratory pathogens. Simultaneous analysis of host and microbe in respiratory samples informs both questions, and is increasingly being applied in studies of the upper and lower airway(22, 46–48). Our work now provides the first fully integrated host/microbe LRTI diagnostic classifier from lower airway mNGS, applicable across pathogen types, thus setting the stage for clinical implementation in the relatively near future.

We envision the approach for LRTI diagnosis by lower airway host/microbe mNGS outlined in this study being used at the time of intubation for critically ill children with acute respiratory failure, as a complement to traditional culture and PCR-based microbiologic testing. Our approach would need to be independently validated and its impact on clinical outcomes would need to be evaluated in a randomized clinical trial before deployment in the hospital. Future work should also examine the trajectory of patient LRTI classification over time, as infection resolves, and how well the classifier might generalize to a similarly large and heterogeneous adult cohort.

## METHODS

### Study cohort

We conducted a secondary analysis of a prospective cohort study of mechanically ventilated children admitted to eight Pediatric Intensive Care units in the National Institute of Child Health and Human Development’s Collaborative Pediatric Critical Care Research Network (CPCCRN) from February 2015 to December 2017(9, 25).

We enrolled children aged 31 days to 18 years who were expected to require mechanical ventilation (MV) via endotracheal tube (ETT) for at least 72 hours. Exclusion criteria included inability to obtain a tracheal aspirate (TA) sample from the subject within 24 hours of intubation; presence of a tracheostomy tube or plans to place one; any condition in which deep tracheal suctioning was contraindicated; previous episode of MV during the hospitalization; family/team lack of commitment to aggressive intensive care as indicated by ‘do not resuscitate’ orders and/or other limitation of care; or previous enrollment into this study. Some patients were ultimately excluded from the present analysis based on sequencing metrics, as described in the following.

Parents or other legal guardians of eligible patients were approached for consent by study-trained staff as soon as possible after intubation. Waiver of consent was granted for TA samples to be obtained from standard-of-care suctioning of the ETT until the parents or guardians could be approached for informed consent.

Prospectively collected clinical data were recorded in a web-based research database maintained by the CPCCRN data coordinating center at the University of Utah.

### Clinical microbiologic diagnostics

Enrolled patients received standard-of-care clinical respiratory microbiologic diagnostics, as ordered by treating clinicians at each study site. These diagnostics included nasopharyngeal (NP) swab respiratory viral testing by multiplex PCR and/or tracheal aspirate (TA) bacterial and fungal semi-quantitative cultures. Clinical diagnostic tests on samples obtained within 48 hours of intubation were included in the analyses. Microbes reported by the clinical laboratory as representing laboratory, skin, or environmental contaminants, or reported as mixed upper respiratory flora, were excluded.

### Adjudication of LRTI status

Adjudication of LRTI status was blinded to the mNGS results and depended on the combination of two elements: i) a retrospective clinical diagnosis made by study-site clinicians, who reviewed all clinical, laboratory and imaging data available at the end of the admission, and ii) any standard-of-care microbiologic diagnostics performed during the admission (nasopharyngeal swab viral PCR and/or TA culture). Following a recently described approach(9, 16, 22), study team physicians ultimately assigned patients into one of four groups: i) Definite, if clinicians made a diagnosis of LRTI and the patient had clinical microbiologic findings; ii) Suspected, if clinicians made a diagnosis of LRTI but there were no microbiologic findings; iii) Indeterminate, if no diagnosis of LRTI was made despite some microbiologic findings; and iv) No Evidence, if clinicians identified a clear non-infectious cause of acute respiratory failure and no clinical or microbiologic suspicion of LRTI arose. We note that comprehensive microbiologic testing was not always performed in the No Evidence group in the absence of clinical suspicion.

### Sample collection, processing and mNGS

Tracheal aspirate (TA) was collected within 24 hours of intubation, mixed 1:1 with DNA/RNA Shield (Zymo) and frozen at −80°C. RNA was extracted from 300µl of patient TA using bead-based lysis and the Allprep DNA/RNA kit (Qiagen), which included a DNase treatment step. RNA was reverse transcribed to generate cDNA, and sequencing library preparation was performed using the NEBNext Ultra II Library Prep Kit. RNA-Seq libraries underwent 150bp paired-end sequencing on an Illumina Novaseq 6000 instrument.

### Host gene expression analysis

Following de-multiplexing, sequencing reads were pseudo-aligned with kallisto(49) (including bias correction) to an index consisting of all transcripts associated with human protein coding and long non-coding RNA genes (ENSEMBL v.99). We excluded samples with less than 500,000 estimated counts associated with transcripts of protein-coding genes. Gene-level counts were generated from the transcript-level abundance estimates using the R package tximport(50), with the scaledTPM method.

Genes were retained for differential expression (DE) analysis if they had at least 10 counts in at least 20% of the samples included in the analysis. DE analyses were performed with the R package limma(51), using quantile normalization and the voom method. P-values were calculated using moderated t-tests, as implemented in limma, and adjusted for multiple hypothesis testing with the Benjamini-Hochberg method. Tests with p < 0.05 were considered significant. Full DE results comparing: i) Definite and No Evidence patients, and ii) Definite patients with any bacterial LRTI and with purely viral LRTI are provided as **Supplemental Data 2**.

Gene set enrichment analyses (GSEA)(52) were performed using the fgseaMultilevel function in the R package fgsea(53), which calculates pathway p-values using an adaptive, multilevel splitting Monte Carlo approach. The analysis was applied to REACTOME(54) pathways with a minimum size of 10 genes and a maximum size of 1,500 genes. All genes from the respective DE analysis were included as input, pre-ranked by the DE test statistic. The gene sets shown in the figures were manually selected to reduce redundancy and highlight diverse biological functions from among those with a Benjamini-Hochberg adjusted p < 0.05. Full GSEA results are provided as **Supplemental Data 3**.

### Classification of LRTI status based on host gene expression features

Genes with at least 10 counts in at least 20% of the Definite (n=117) and No Evidence (n=50) patients were used as input for the host-based LRTI classification (n=13,323). We applied a variance-stabilizing transformation to the gene counts, as implemented in the R package DESeq2(55).

We implemented a 5-fold cross-validation procedure such that in each train/test split, we

1. used lasso logistic regression on the samples in the training folds for feature (gene) selection,
2. trained a random forest classifier on the samples in the training folds using only the selected features, and iii) applied the random forest classifier to the samples in the test fold to obtain an out-of-fold host probability of LRTI. We required at least 9 No Evidence patients in each of the folds to ensure sufficient negative samples in each test set.

Simple lasso logistic regression was fit using the cv.glmnet(family=’binomial’) function from the R package glmnet(56), leaving all other parameters at their defaults. We used the 1se criterion for selecting the tuning parameter, which picks the sparsest value of the tuning parameter that lies within 1 standard error of the optimum. When evaluating test error, we selected the tuning parameter via nested cross-validation within the training set only.

Random forest was implemented using the R package randomForest(57). We used 10,000 trees and left all parameters at their defaults.

The area under the receiver operating characteristic curve (AUC) for each test fold was calculated using the R package pROC(58) with default behavior. Sensitivity and specificity were calculated using a pre-determined 50% out-of-fold LRTI probability threshold.

### Detection of microbes by mNGS and background filtering

We processed patient TA samples alongside water controls through the open-source CZ-ID (formerly called IDSeq) metagenomic analysis pipeline(59). The pipeline performs subtractive alignment of the human genome and then reference-based alignment of the remaining reads at both the nucleotide and amino acid level against sequences in the National Center for Biotechnology Information (NCBI) nucleotide (NT) and non-redundant (NR) databases, respectively. This is followed by assembly of the reads matching each taxon. Taxa with ≥5 read counts in the NT alignment and an average assembly nucleotide alignment ≥70bp were retained for downstream analysis.

Water controls enabled estimation of the number of background reads expected for each taxon, as previously described(9, 47). This was done by modeling the number of background reads as a negative binomial distribution with mean and dispersion fitted on the water controls. For each batch (sequencing run) and taxon, we estimated the mean parameter of the negative binomial distribution by averaging the read counts across the water controls after normalizing by the total non-host reads, slightly regularizing this estimate by including the global average (across all batches) as an additional sample. We estimated a single dispersion parameter across all taxa and batches using the functions glm.nb() and theta.md() from the R package MASS(60). Taxa were then tested for whether they exceeded the count expected from the background distribution, and a Benjamini-Hochberg adjustment was applied to all tests performed in the same sample. Taxa were considered present in a sample if they achieved an adjusted p < 0.05.

Any virus with known ability to cause LRTI, based on a previously conducted literature curation(22), that was present in a patient sample after background filtering was considered a probable pathogen.

### Rules-based model (RBM)

For bacteria and fungi that were present after background filtering, we additionally applied slightly adapted from its previously published version(22). Application of the RBM in each sample involved the following steps:

1. We retained only the most abundant bacterial/fungal species from each genus. In case a less abundant species in the genus had known ability to cause LRTI, based on a previously conducted literature curation(22), we retained it too.
2. We then ranked the retained species from greatest to least overall abundance in the sample and limited, at most, to the top 15.
3. The largest drop in abundance between the ranked species in the sample was identified.
4. If any species above the largest drop in abundance had known ability to cause LRTI, it was deemed a potential pathogen.

### Analysis of microbiome diversity

The Shannon diversity index was calculated using either all viral and bacterial taxa, or only bacterial taxa, that were present after background filtering using the R package Vegan(61). Two-sided Mann-Whitney tests with Bonferroni correction were used to evaluate statistical significance of group differences. Tests with p < 0.05 were considered significant.

### Classification of LRTI status based on integration of host and microbial features

For the integrated host/microbe LRTI classifier, we fit a logistic regression model on the following features: i) the host LRTI probability; ii) the summed abundance, measured in reads-per-million (rpM), of any pathogenic viruses present after background filtering (‘viral score’); and iii) the proportion of any potentially pathogenic bacteria/fungi identified by the RBM out of all non-host read counts (‘bacterial score’). To avoid any leakage from the test set affecting the host probabilities, we always used the out-of-bag ‘votes’ from the host random forest classifier as the host probabilities of the training samples.

Before fitting the integrated classifier, we applied transformations to all three features. A logistic (log-odds) transformation was applied to the host probabilities: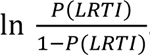. To facilitate this transformation, we first slightly regularized the host probabilities and their complementary probabilities away from 0 and 1 by a quantity inversely proportional to the number of random forest trees used in the host classifier (*RF_trees_* = 10,000):

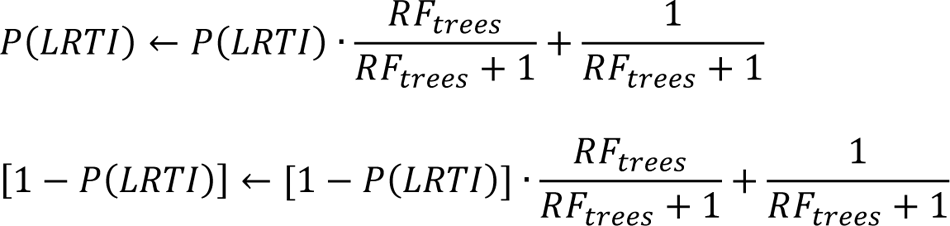

For the viral/bacterial scores, we applied a log_10_ transformation. In order to avoid taking the log of 0, we added a small uniform quantity to the scores of all the samples, which was calculated by taking the minimum non-zero viral or bacterial score, respectively, in the corresponding training set and dividing it by 10.

Performance of the integrated classifier was evaluated on the Definite and No Evidence patients using 5-fold cross-validation, with the same train/test splits and the same per-split host classifiers as in the host-only cross-validation. The area under the receiver operating characteristic curve (AUC) for each test fold was calculated using the R package pROC(58) with default behavior. Sensitivity and specificity were calculated using a pre-determined 50% out-of-fold LRTI probability threshold.

The integrated classifier was then trained on all the Definite and No Evidence patients and applied to the Suspected and Indeterminate patients.

#### Statistics

This study implemented a 5-fold cross-validation scheme to develop and evaluate performance of a binary classifier using samples with presumed known labels. Algorithms used in the classification procedure included logistic regression and random forest, which generate a probabilistic classification output. The area under the receiver operating characteristic curve, as well as sensitivity and specificity at a pre-determined probability threshold of 0.5, were used as performance metrics. Detailed descriptions of each statistical analysis are included in the corresponding sections of the Methods and in the figure and table legends.

#### Study approval

The original cohort study was approved by the Collaborative Pediatric Critical Care Research IRB at the University of Utah (protocol #00088656). Informed consent was obtained from parents or other legal guardians, which included permission for collected specimens and data to be used in future studies.

#### Data and code availability

Raw FASTQ files are protected due to patient privacy concerns. Processed host gene counts are available in the NCBI Gene Expression Omnibus (GEO) database under accession <u>GSE212532</u>. FASTQ files containing non-host reads identified by the CZ-ID pipeline, following subtraction of reads aligning to the human genome, are available in the NCBI Sequence Read Archive (SRA) database under BioProject accession <u>PRJNA875913</u>. All data, code, and results related to development and validation of the mNGS classifier, including the microbial taxon counts, are available at: https://github.com/eranmick/pediatric-mNGS-LRTI-classifier. Supplementary data files are provided with this publication.

## Author contributions

EM, AT, JK, KLK, JLD, PMM, and CRL contributed to study conception and overall design. SC, AL, MT, AMD, NN, and CRL oversaw or performed sample processing, library preparation, and sequencing. EM, AT, JK, CMO, KMW, VS, ABM, BDW, LA, and CRL oversaw or performed collation and annotation of patient metadata. CMO, ABM, TC, PMM, and CRL contributed to patient clinical adjudication. JK designed the underlying cross-validation scheme. EM, AT, JK, and CRL performed all data analyses and data visualizations. KLK contributed to implementation of the rules-based model. LA contributed to project administration and coordination. EM, AT, PMM, and CRL wrote the manuscript with input from all authors. ML, ABM, EAFS, TC, and BDW provided advice and feedback throughout the study. PMM and CRL jointly supervised the study. EM, AT, and JK are listed as co-first authors due to equal contribution to the study, with the order of appearance determined alphabetically by last name except in the case of JK who had departed the group by the time of manuscript preparation.

## Data Availability

Processed gene counts are available in the NCBI Gene Expression Omnibus (GEO) database under accession GSE212532.

## Acknowledgements

We thank all subjects and their families for participating in this study. We acknowledge the contributions of Tammara L. Jenkins, MSN, RN, and Robert F. Tamburro, MD, from Eunice Kennedy Shriver National Institute of Child Health and Human Development, Bethesda, MD. The following is a summary of study sites where patients were enrolled, principal investigators (PI), co-investigators (CI) and research coordinators (RC). Children’s Hospital of Colorado, Aurora, CO: Peter Mourani (PI); Todd Carpenter (CI); Yamila Sierra (RC); Katheryn Malone (RC), Diane Ladell (RC); Kimberly Ralston (RC); Kevin Van (RC). Children’s Hospital of Michigan, Detroit, MI: Kathleen L. Meert (PI); Sabrina Heidemann (CI); Ann Pawluszka (RC); Melanie Lulic (RC). Children’s Hospital of Philadelphia, Philadelphia, PA: Robert A Berg (PI); Athena Zuppa (CI); Carolann Twelves (RC); Mary Ann DiLiberto (RC). Children’s National Medical Center, Washington, DC: Murray Pollack (PI); David Wessel (PI); Randall Burd (CI); Elyse Tomanio (RC); Diane Hession (RC); Ashley Wolfe (RC). Nationwide Children’s Hospital, Columbus, OH: Mark Hall (PI); Andrew Yates (CI); Lisa Steele (RC); Maggie Flowers (RC); Josey Hensley (RC). Mattel Children’s Hospital, University of California Los Angeles, Los Angeles, CA: Anil Sapru (PI); Rick Harrison (CI), Neda Ashtari (RC); Anna Ratiu (RC). Children’s Hospital of Pittsburgh, University of Pittsburgh Medical Center, Pittsburgh, PA: Joe Carcillo (PI); Ericka Fink (CI); Leighann Koch (RC); Alan Abraham (RC). Benioff Children’s Hospital, University of California, San Francisco, San Francisco, CA: Patrick McQuillen (PI); Anne McKenzie (RC); Yensy Zetino (RC). We also acknowledge the support of the University of Utah Data Coordinating Center, Salt Lake City, Utah: Mike Dean (PI); Richard Holubkov (PI), Juhee Peterson, Melissa Bolton, Whit Coleman, and Stephanie Dorton. This study was supported in part by the following cooperative agreements from the Eunice Kennedy Shriver National Institute of Child Health and Human Development and the National Heart, Lung and Blood Institute: UG1HD083171 (Dr. Mourani), 1R01HL124103 (Drs. Mourani and Sontag), UG1HD049983 (Dr. Carcillo), UG01HD049934 (Drs. Reeder, Locandro, and Dean), UG1HD083170 (Dr. Hall), UG1HD050096 (Dr. Meert), UG1HD63108 (Dr. Zuppa), UG1HD083116 (Dr. Sapru), UG1HD083166 (Dr. McQuillen), UG1HD049981 (Dr. Pollack), K23HL138461-01A1 and 5R01HL155418-02 (Dr. Langelier). The study was also supported by funding from the Chan Zuckerberg Biohub (Dr. Langelier). Study sponsors were not involved in study design; in the collection, analysis, and interpretation of data; in the writing of the report; and in the decision to submit the report for publication.

## Conflict of interest statement

EM, AT, JK, KLK, PMM, and CRL are listed as inventors on a patent application (63/381,156) related to the diagnosis of lower respiratory tract infections filed by the University of California, San Francisco and the Chan Zuckerberg Biohub.

## Supplemental Figures

**Supplemental Figure 1:**
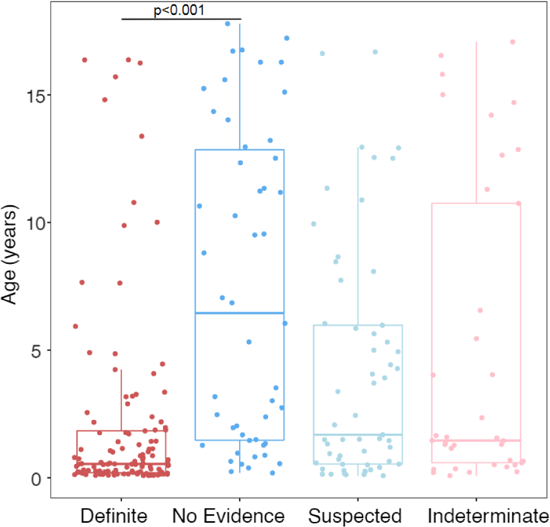
Age distribution across the four LRTI status groups. P-value for the comparison between Definite and No Evidence patients was calculated using a Mann-Whitney test.

**Supplemental Figure 2:**
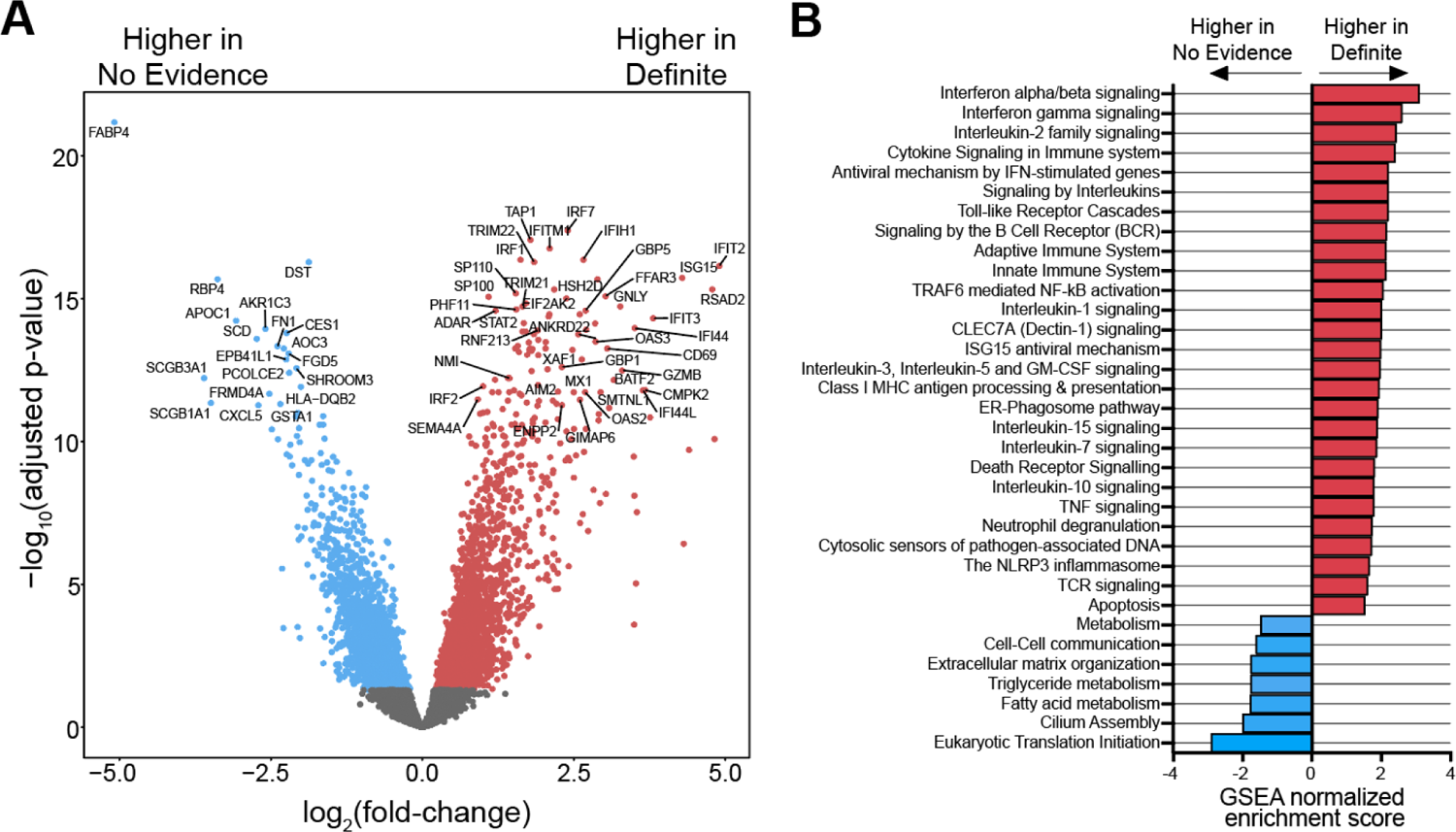
**A)** Volcano plot highlighting genes differentially expressed (DE) between Definite and No Evidence patients. Colored genes reached statistical significance (adjusted p-value < 0.05). **B)** Normalized enrichment scores of selected REACTOME pathways that reached statistical significance (adjusted p-value < 0.05) in the GSEA using DE genes between Definite and No Evidence patients.

**Supplemental Figure 3:**
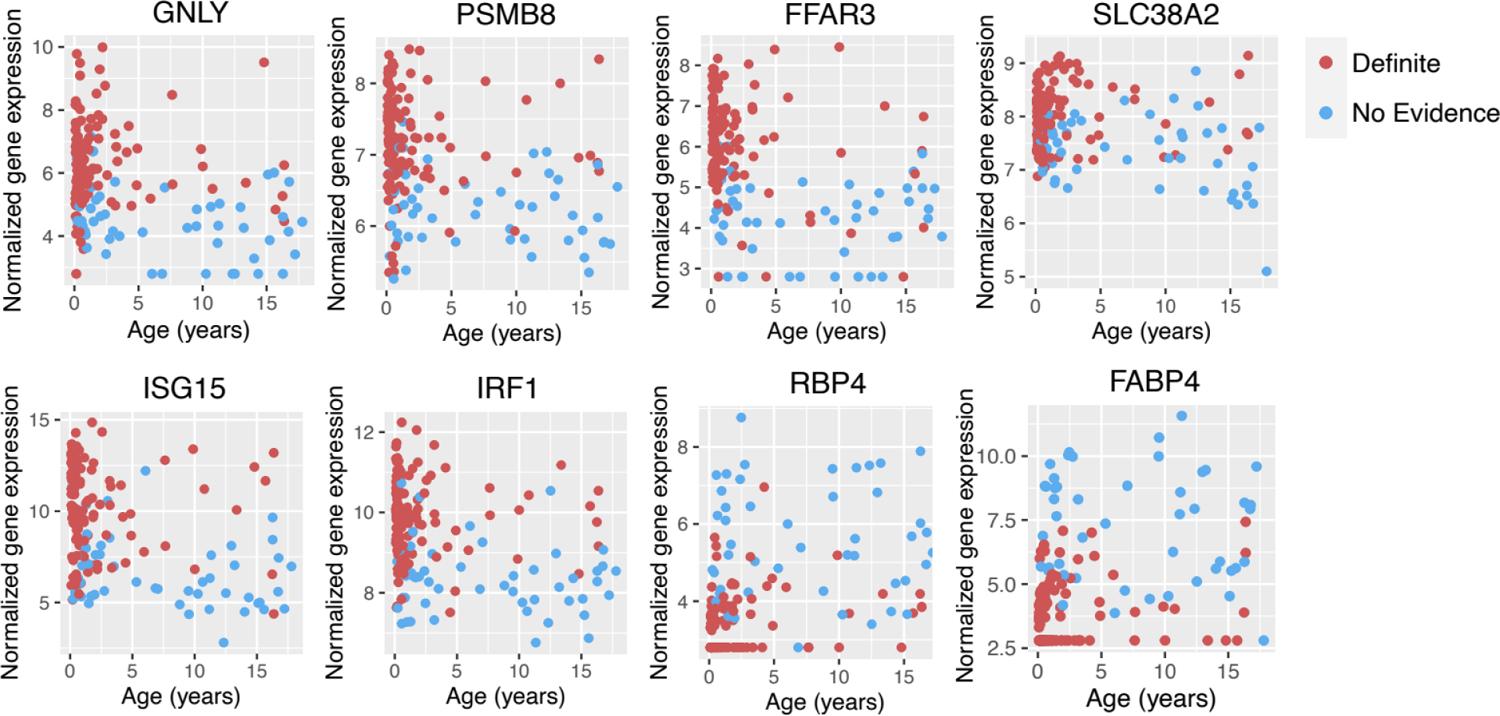
Expression of eight host classifier genes as a function of age in Definite (red) and No Evidence (blue) patients.

**Supplemental Figure 4:**
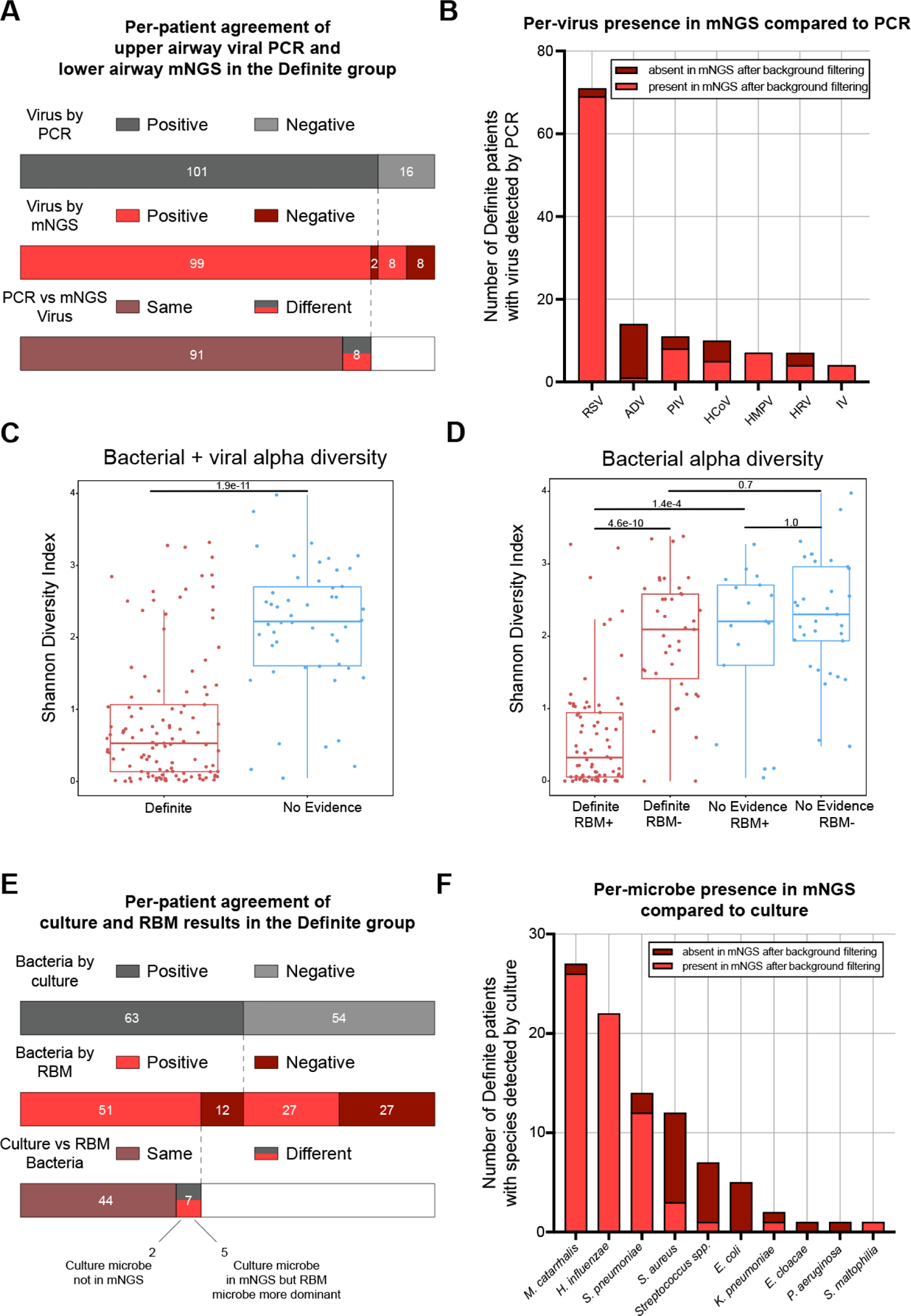
**A)** Diagram depicting the agreement at the patient level between clinical upper respiratory PCR viral testing and lower airway mNGS detection after background filtering in the Definite group. Agreement between the two methods in a patient was defined as at least one virus identified by both. **B)** Bar plot showing the number of cases of each virus detected by clinical upper respiratory PCR testing and the proportion that was also present by mNGS after background filtering. RSV, respiratory syncytial virus; ADV, adenovirus; PIV, parainfluenza virus; HCoV, human coronavirus; HMPV, human metapneumovirus; HRV, human rhinovirus; IV, influenza virus. **C)** Boxplots of bacterial+viral microbiome alpha diversity, measured by the Shannon index, in Definite and No Evidence patients. Horizontal lines denote the median, box hinges represent the interquartile range (IQR), and whiskers extend to the most extreme value no greater than 1.5*IQR from the hinges. **D)** Boxplots of bacterial-only alpha diversity measured by the Shannon index. Definite and No Evidence patients are split by whether a potential pathogen was identified by the RBM. P-values in C) and D) were calculated by a Mann-Whitney test with Bonferroni correction. **E)** Diagram depicting the agreement at the patient level between clinical culture and the results of the RBM in the Definite group. Agreement between the two methods in a patient was defined as at least one species identified by both. **F)**Bar plot showing the number of cases of each species detected by clinical culture and the proportion that was also present by mNGS after background filtering. *Streptococcus spp.*, *Streptococcus* species other than *S. pneumoniae*.

**Supplemental Figure 5:**
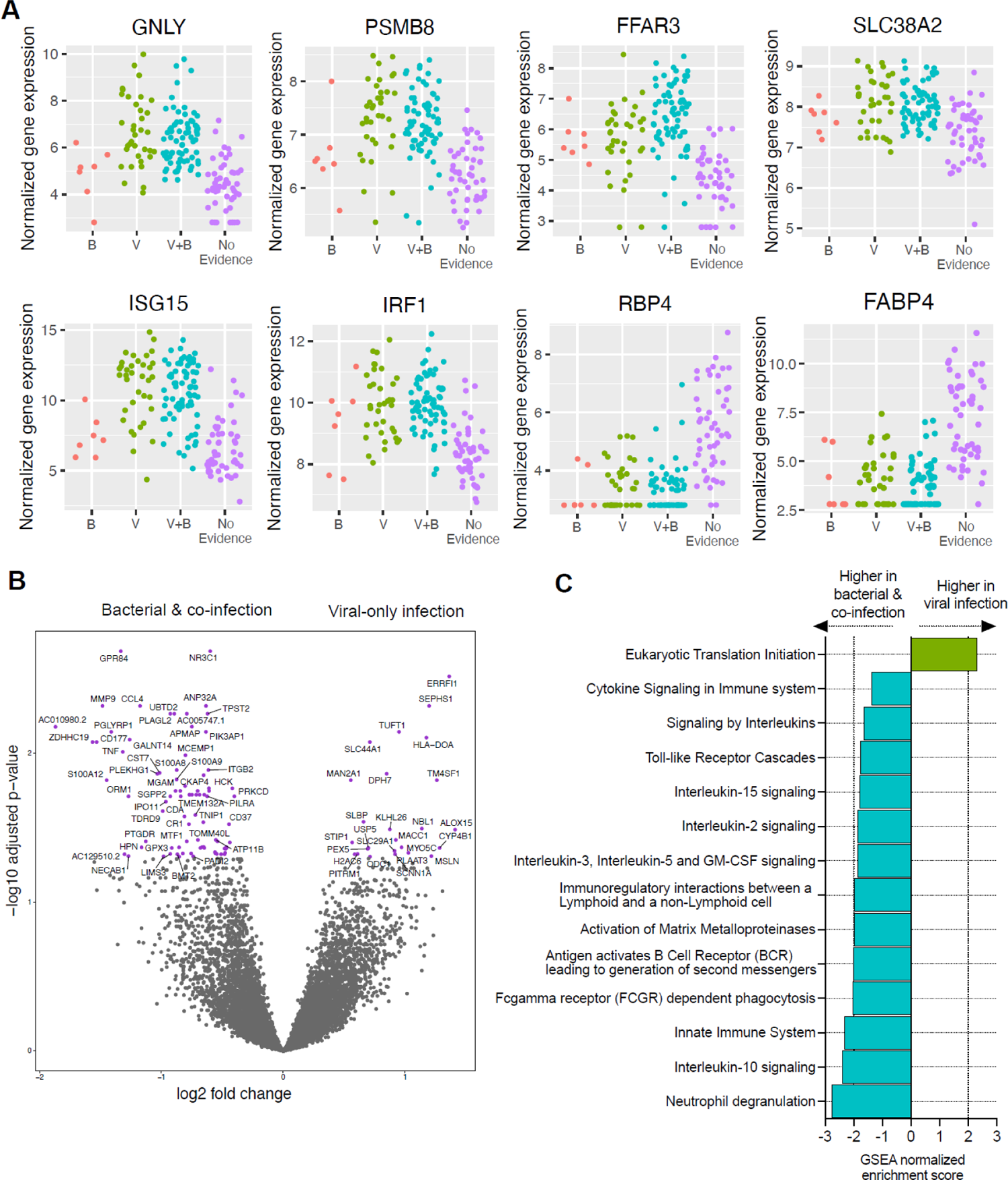
**A)** Expression of eight host classifier genes in Definite patients with only bacterial pathogens identified by the RBM (B; n=7), only viral pathogens detected by mNGS (V; N=36), viral and bacterial pathogens (V+B; n=71), and the No Evidence patients (n=50) for comparison. Three patients from the Definite group are not shown because they did not have any pathogens identified by mNGS. One No Evidence sample in the plot of *SLC38A2* was omitted since it was an extreme outlier. **B)** Volcano plot highlighting genes differentially expressed (DE) between Definite patients with any bacterial infection (bacterial-only + co-infection) and viral-only infection. Genes colored in purple reached statistical significance (adjusted p-value < 0.05). **C)** Normalized enrichment scores of selected REACTOME pathways that reached statistical significance (adjusted p-value < 0.05) in the GSEA using the DE genes.

**Supplemental Figure 6:**
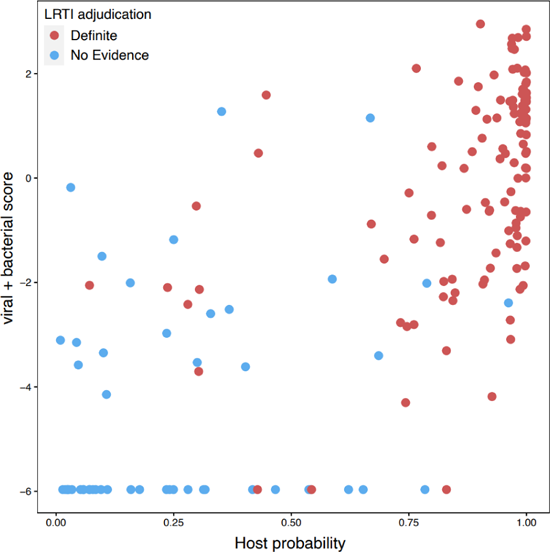
Scatterplot of the host LRTI probability (x-axis) and the sum of the log_10_-transformed microbial scores (y-axis) in the Definite and No Evidence patients.

## Supplemental Tables

**Supplemental Table 1:** Host genes selected by lasso logistic regression in each of the 5 cross-validation train/test splits, and the area under the receiver operating characteristic curve (AUC) of a random forest classifier using the selected genes.

| Test fold | Gene ID | Gene symbol | Regression coefficient | AUC |
| --- | --- | --- | --- | --- |
| 1 | (Intercept) | NA | -2.0280997 | 0.996 |
| 1 | ENSG00000115523 | *GNLY* | 0.40435424 |  |
| 1 | ENSG00000114737 | *CISH* | 0.29736499 |  |
| 1 | ENSG00000185897 | *FFAR3* | 0.2334893 |  |
| 1 | ENSG00000134294 | *SLC38A2* | 0.23225034 |  |
| 1 | ENSG00000204264 | *PSMB8* | 0.14737796 |  |
| 1 | ENSG00000133101 | *CCNA1* | 0.13698008 |  |
| 1 | ENSG00000125347 | *IRF1* | 0.08788657 |  |
| 1 | ENSG00000152766 | *ANKRD22* | 0.01618487 |  |
| 1 | ENSG00000103356 | *EARS2* | -0.005208 |  |
| 1 | ENSG00000272168 | *CASC15* | -0.0129449 |  |
| 1 | ENSG00000163710 | *PCOLCE2* | -0.0141204 |  |
| 1 | ENSG00000196139 | *AKR1C3* | -0.0145091 |  |
| 1 | ENSG00000163735 | *CXCL5* | -0.0202633 |  |
| 1 | ENSG00000164631 | *ZNF12* | -0.0296153 |  |
| 1 | ENSG00000162929 | *KIAA1841* | -0.031873 |  |
| 1 | ENSG00000102962 | *CCL22* | -0.0470626 |  |
| 1 | ENSG00000196305 | *IARS1* | -0.0541444 |  |
| 1 | ENSG00000259094 | *AC013457.1* | -0.0642868 |  |
| 1 | ENSG00000203865 | *ATP1A1-AS1* | -0.0785439 |  |
| 1 | ENSG00000115414 | *FN1* | -0.0963792 |  |
| 1 | ENSG00000132300 | *PTCD3* | -0.1094826 |  |
| 1 | ENSG00000273173 | *SNURF* | -0.1236364 |  |
| 1 | ENSG00000138207 | *RBP4* | -0.1867265 |  |
| 1 | ENSG00000170323 | *FABP4* | -0.2260672 |  |
| 1 | ENSG00000182141 | *ZNF708* | -0.2407744 |  |
| 2 | (Intercept) | NA | -0.4183313 | 0.953 |
| 2 | ENSG00000134294 | *SLC38A2* | 0.21115084 |  |
| 2 | ENSG00000185897 | *FFAR3* | 0.17219249 |  |
| 2 | ENSG00000115523 | *GNLY* | 0.15221918 |  |
| 2 | ENSG00000204264 | *PSMB8* | 0.0974774 |  |
| 2 | ENSG00000152766 | *ANKRD22* | 0.06800369 |  |
| 2 | ENSG00000272821 | *U62317.2* | 0.05245633 |  |
| 2 | ENSG00000187608 | *ISG15* | 0.03953433 |  |
| 2 | ENSG00000162929 | *KIAA1841* | -0.0224986 |  |
| 2 | ENSG00000196139 | *AKR1C3* | -0.0991982 |  |
| 2 | ENSG00000170323 | *FABP4* | -0.1720699 |  |
| 2 | ENSG00000253729 | *PRKDC* | -0.2010562 |  |
| 2 | ENSG00000138207 | *RBP4* | -0.2940594 |  |
| 3 | (Intercept) | NA | 0.27758103 | 0.986 |
| 3 | ENSG00000185507 | *IRF7* | 0.29760005 |  |
| 3 | ENSG00000133101 | *CCNA1* | 0.22848885 |  |
| 3 | ENSG00000115523 | *GNLY* | 0.20275662 |  |
| 3 | ENSG00000185897 | *FFAR3* | 0.09319602 |  |
| 3 | ENSG00000134294 | *SLC38A2* | 0.05523801 |  |
| 3 | ENSG00000272821 | *U62317.2* | 0.03959668 |  |
| 3 | ENSG00000196189 | *SEMA4A* | 0.03111383 |  |
| 3 | ENSG00000136231 | *IGF2BP3* | 0.01793655 |  |
| 3 | ENSG00000114737 | *CISH* | 0.01066616 |  |
| 3 | ENSG00000117143 | *UAP1* | -0.0043278 |  |
| 3 | ENSG00000163735 | *CXCL5* | -0.0087013 |  |
| 3 | ENSG00000113068 | *PFDN1* | -0.0087115 |  |
| 3 | ENSG00000149021 | *SCGB1A1* | -0.0088169 |  |
| 3 | ENSG00000232629 | *HLA-DQB2* | -0.0158939 |  |
| 3 | ENSG00000164631 | *ZNF12* | -0.0351792 |  |
| 3 | ENSG00000273173 | *SNURF* | -0.0502438 |  |
| 3 | ENSG00000196139 | *AKR1C3* | -0.0532949 |  |
| 3 | ENSG00000170324 | *FRMPD2* | -0.0607793 |  |
| 3 | ENSG00000259094 | *AC013457.1* | -0.0637096 |  |
| 3 | ENSG00000272660 | *AC090425.2* | -0.1024448 |  |
| 3 | ENSG00000008226 | *DLEC1* | -0.1276673 |  |
| 3 | ENSG00000272168 | *CASC15* | -0.1499889 |  |
| 3 | ENSG00000170323 | *FABP4* | -0.4076148 |  |
| 4 | (Intercept) | NA | -5.181784 | 0.954 |
| 4 | ENSG00000175073 | *VCPIP1* | 0.50212823 |  |
| 4 | ENSG00000115523 | *GNLY* | 0.23413097 |  |
| 4 | ENSG00000135604 | *STX11* | 0.23226792 |  |
| 4 | ENSG00000168394 | *TAP1* | 0.13854581 |  |
| 4 | ENSG00000185897 | *FFAR3* | 0.13639215 |  |
| 4 | ENSG00000185885 | *IFITM1* | 0.07866599 |  |
| 4 | ENSG00000133106 | *EPSTI1* | 0.05899777 |  |
| 4 | ENSG00000158769 | *F11R* | -0.0055638 |  |
| 4 | ENSG00000163710 | *PCOLCE2* | -0.0206461 |  |
| 4 | ENSG00000149021 | *SCGB1A1* | -0.0263191 |  |
| 4 | ENSG00000182141 | *ZNF708* | -0.1006603 |  |
| 4 | ENSG00000138207 | *RBP4* | -0.1413799 |  |
| 4 | ENSG00000170323 | *FABP4* | -0.2005039 |  |
| 5 | (Intercept) | NA | 0.5726104 | 0.967 |
| 5 | ENSG00000204264 | *PSMB8* | 0.27278452 |  |
| 5 | ENSG00000115523 | *GNLY* | 0.0762971 |  |
| 5 | ENSG00000175073 | *VCPIP1* | 0.05579582 |  |
| 5 | ENSG00000185897 | *FFAR3* | 0.04184826 |  |
| 5 | ENSG00000188820 | *CALHM6* | 0.03110363 |  |
| 5 | ENSG00000133106 | *EPSTI1* | 0.02410902 |  |
| 5 | ENSG00000187608 | *ISG15* | 0.00687972 |  |
| 5 | ENSG00000163710 | *PCOLCE2* | -0.0191247 |  |
| 5 | ENSG00000151914 | *DST* | -0.029031 |  |
| 5 | ENSG00000163735 | *CXCL5* | -0.1149234 |  |
| 5 | ENSG00000170323 | *FABP4* | -0.389078 |  |

**Supplemental Table 2:**
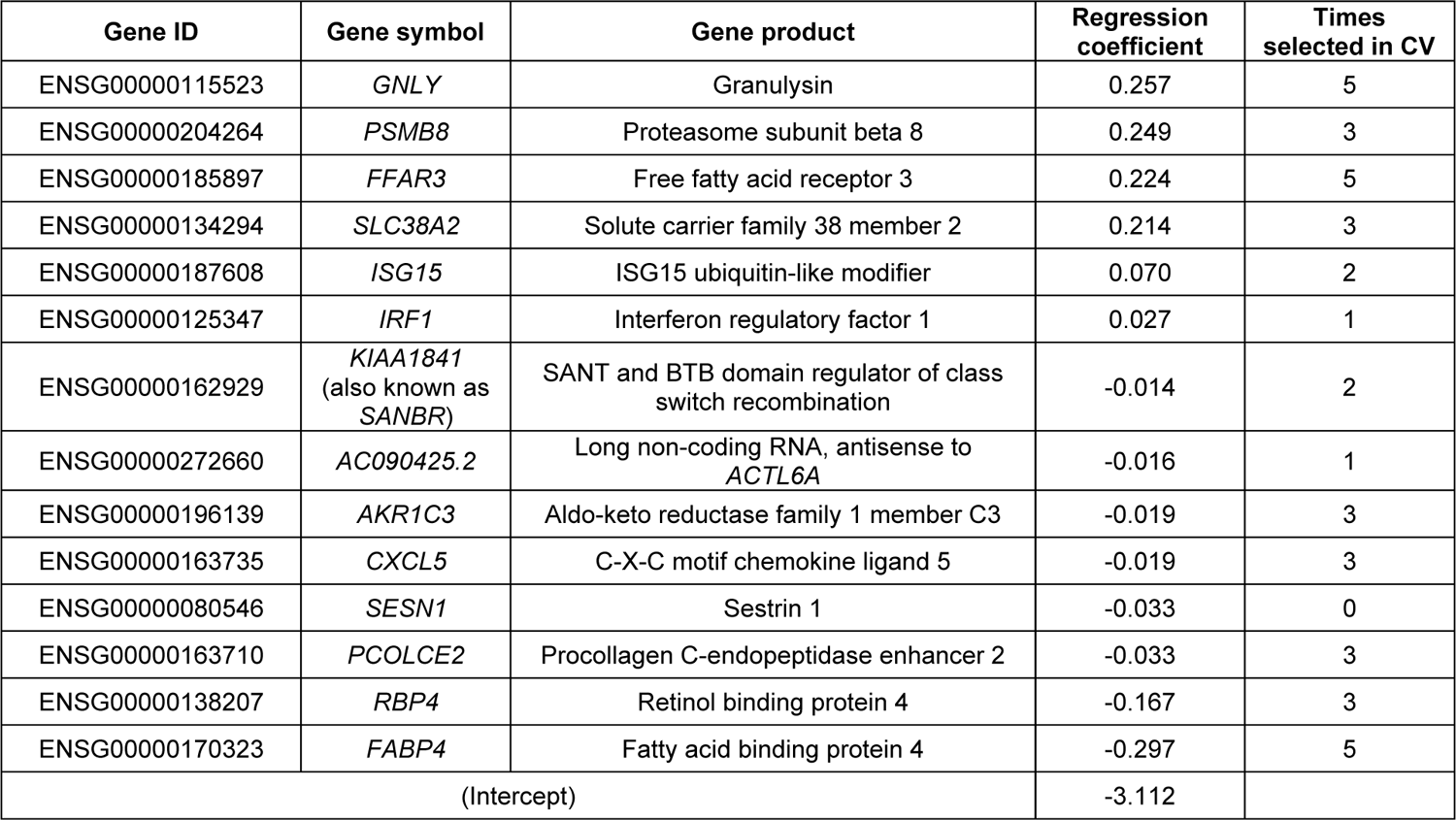
Genes selected for the final host classifier by lasso logistic regression applied to all the Definite and No Evidence patients, with their regression coefficients. The number of times the gene was selected across the 5 cross-validation (CV) splits is also indicated.

**Supplemental Table 3:** Differential expression results for the 14 final classifier genes comparing: **A)** No Evidence patients under four years old (n=23; median age 1.3 years) versus over four years old (n=27; median age 12.5), and **B)** Definite patients under four years old (n=100; median age 0.4) versus No Evidence patients under four years old (n=23; median age 1.3).

**A**
| Gene symbol | Log<sub>2</sub> fold-change | P-value | Adjusted P-value |
| --- | --- | --- | --- |
| *GNLY* | -1.14 | 0.01 | 0.72 |
| *PSMB8* | -0.33 | 0.21 | 0.82 |
| *FFAR3* | -0.43 | 0.33 | 0.87 |
| *SLC38A2* | -0.57 | 0.07 | 0.75 |
| *ISG15* | -1.68 | 0.01 | 0.72 |
| *IRF1* | -0.30 | 0.25 | 0.83 |
| *KIAA1841* | 0.14 | 0.59 | 0.94 |
| *AC090425.2* | 1.48 | 0.07 | 0.76 |
| *AKR1C3* | 0.81 | 0.10 | 0.76 |
| *CXCL5* | 0.24 | 0.67 | 0.95 |
| *SESN1* | 0.27 | 0.47 | 0.91 |
| *PCOLCE2* | 0.54 | 0.21 | 0.81 |
| *RBP4* | -0.04 | 0.95 | 0.99 |
| *FABP4* | -0.56 | 0.45 | 0.90 |

**B**
| Gene symbol | Log<sub>2</sub> fold-change | P-value | Adjusted P-value |
| --- | --- | --- | --- |
| *GNLY* | 2.73 | 2.11E-08 | 1.46E-06 |
| *PSMB8* | 1.11 | 7.78E-08 | 4.22E-06 |
| *FFAR3* | 2.93 | 9.70E-12 | 4.25E-09 |
| *SLC38A2* | 0.60 | 2.50E-05 | 4.33E-04 |
| *ISG15* | 3.49 | 3.71E-09 | 3.89E-07 |
| *IRF1* | 1.52 | 5.18E-12 | 2.88E-09 |
| *KIAA1841* | -0.89 | 2.84E-05 | 4.76E-04 |
| *AC090425.2* | -0.18 | 7.50E-01 | 8.56E-01 |
| *AKR1C3* | -2.48 | 2.24E-12 | 1.42E-09 |
| *CXCL5* | -2.63 | 4.62E-09 | 4.52E-07 |
| *SESN1* | -0.53 | 4.92E-02 | 1.38E-01 |
| *PCOLCE2* | -2.06 | 2.60E-09 | 2.93E-07 |
| *RBP4* | -3.64 | 1.51E-17 | 1.00E-13 |
| *FABP4* | -5.54 | 4.02E-26 | 5.36E-22 |

**Supplemental Table 4:** Comparison of mNGS viral detection in TA samples with PCR viral detection in nasopharyngeal (NP) swabs or in the same TA samples in a subset of patients.

| Definite patients with matched NP swab and TA viral PCR testing (n=21) | Agreement of mNGS with NP swab PCR | Concordance of mNGS and NP swab PCR | Agreement of mNGS with TA PCR | Concordance of mNGS and TA PCR |
| --- | --- | --- | --- | --- |
| All viruses | 22/34 = 64.7% | 22/37 = 59.5% | 23/24 = 95.8% | 23/26 = 88.5% |
| Respiratory syncytial virus | 11/12 = 91.7% | 11/12 = 91.7% | 10/10 = 100% | 10/11 = 90.9% |
| Rhinovirus | 4/7 = 57.1% | 4/10 = 40% | 6/6 = 100% | 6/7 = 85.7% |
| Adenovirus | 0/6 = 0% | 0/6 = 0% | 0/0 = 100% | 0/0 = 100% |
| Coronavirus | 2/3 = 66.7% | 2/3 = 66.7% | 2/2 = 100% | 2/2 = 100% |
| Human metapneumovirus | 3/3 = 100% | 3/3 = 100% | 3/3 = 100% | 3/3 = 100% |
| Parainfluenza virus | 2/3 = 66.7% | 2/3 = 66.7% | 2/3 = 66.7% | 2/3 = 66.7% |

Agreement reflects the number of viruses detected by mNGS out of the total number of viruses detected by PCR.
Concordance reflects the number of viruses detected by both mNGS and PCR out of the total number of viruses detected by at least one method.

**Supplemental Table 5:**
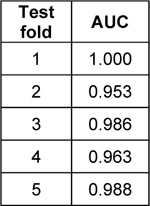
Per-fold area under the curve (AUC) values for the integrated host/microbe logistic regression classifier.

| Test fold | AUC |
| --- | --- |
| 1 | 1.000 |
| 2 | 0.953 |
| 3 | 0.986 |
| 4 | 0.963 |
| 5 | 0.988 |

**Supplemental Table 6:** **A)** mNGS and clinical microbiology results for the Definite and No Evidence patients whose integrated LRTI classification was inconsistent with their adjudication. **B)** Primary diagnoses of the No Evidence patients whose integrated LRTI classification was inconsistent with their adjudication.

**A**
| Patient | LRTI adjudication | Host P(LRTI) | Clinical microbiology results | mNGS viruses | Viral ΣrpM | mNGS RBM hits | Dominance of RBM hits | Integ. P(LRTI) |
| --- | --- | --- | --- | --- | --- | --- | --- | --- |
| P2 | Definite | 0.30 | *S. aureus*,<br>HRV |  | 0 | *S. aureus* | 0.62 | 0.10 |
| P3 | Definite | 0.30 | HRV,<br>ADV | HRV C | 2.15 |  | 0 | 0.22 |
| P6 | Definite | 0.24 | PIV,<br>HCoV | PIV 4,<br>HCoV NL63 | 87.61 |  | 0 | 0.29 |
| P124 | Definite | 0.28 | HCoV,<br>*S. aureus*,<br>*S. viridans* | HCoV 229E | 41.58 |  | 0 | 0.41 |
| P185 | Definite | 0.07 | *S. maltophilia*,<br>*S. pneumoniae*,<br>*K. pneumoniae* |  | 0 | *S. maltophilia*,<br>*S. pneumoniae* | 0.75 | 0.01 |
| P189 | Definite | 0.30 | *S. aureus* | CMV | 0.38 | *S. aureus* | 0.75 | 0.43 |
| P195 | Definite | 0.54 | *S. viridans*,<br>*E. coli* |  | 0 |  | 0 | 0.23 |
| P218 | Definite | 0.43 | *M. catarrhalis* |  | 0 |  | 0 | 0.16 |
| P1 | No Evidence | 0.67 | No testing performed | HCoV NL63 | 44.94 | *P. aeruginosa* | 0.31 | 0.95 |
| P30 | No Evidence | 0.35 | No culture performed,<br>negative PCR | HRV C | 22.15 | *M. catarrhalis* | 0.85 | 0.68 |
| P75 | No Evidence | 0.96 | No culture performed,<br>negative PCR | RSV | 44.53 |  | 0 | 1.00 |
| P166 | No Evidence | 0.79 | No testing performed |  | 0 | *S. aureus* | 0.82 | 0.92 |
| P175 | No Evidence | 0.69 | No testing performed | HCoV NL63 | 4.33 |  | 0 | 0.78 |
| P250 | No Evidence | 0.78 | Negative culture,<br>no PCR performed |  | 0 |  | 0 | 0.62 |

| Patient | LRTI adjudication | Primary diagnosis |
| --- | --- | --- |
| P1 | No Evidence | Neurological |
| P30 | No Evidence | Trauma |
| P75 | No Evidence | Non-infectious respiratory distress |
| P166 | No Evidence | Ingestion (drug/toxin) |
| P175 | No Evidence | Ingestion (drug/toxin) |
| P250 | No Evidence | Seizures |

## Supplemental Data Files

**Supplemental Data File 1.** Basic sample metadata.

**Supplemental Data File 2.** Differential expression (DE) analyses between: i) Definite and No Evidence patients; ii) Definite patients with any bacterial LRTI and with purely viral LRTI.

**Supplemental Data File 3.** Gene set enrichment analysis (GSEA) results from the DE between: i) Definite and No Evidence patients; ii) Definite patients with any bacterial LRTI and with purely viral LRTI.

**Supplemental Data File 4.** Pathogens identified in Definite patients by clinical testing and by mNGS.

## Notes

### Competing Interest Statement

The authors have declared no competing interest.

### Funding Statement

NHLBI, Chan Zuckerberg Biohub

### Author Declarations

Multi-institutional study approved by the single Collaborative Pediatric Critical Care Research IRB at the University of Utah (protocol #00088656).

### Summary of Updates

Updates to data sharing, text, figures. No new analyses.

